# Plant-based whole-food diets are feasible during autologous stem cell transplantation and are associated with dose-dependent microbiome modulation: Results from a pilot clinical trial

**DOI:** 10.64898/2026.02.02.26345403

**Authors:** Katherine Ueland, Taara Elahi, Mary Rasmussen, Alex E. Wolfe, Hayley Purcell, Saranya Rani Chakka, Mercy Mirimo-Martinez, Heather Persinger, Karen Johnson, Alanna Boynton, Kerry McMillen, Mariia Byelykh, Melinda A. Biernacki, Albert C. Yeh, Naveed Ali, Shivaprasad Manjappa, Natalie Wuliji, David N. Fredricks, Marie Bleakley, Leona A. Holmberg, Jeannette M. Schenk, Daniel Raftery, Jing Ma, Geoffrey R. Hill, Marian L. Neuhouser, Stephanie J. Lee, Kate A. Markey

## Abstract

Plant-based dietary strategies may offer a tractable approach to mitigating microbiome disruption and improving outcomes in patients undergoing autologous hematopoietic cell transplantation (auto-HCT) for multiple myeloma, a population in whom intestinal dysbiosis has been linked to infectious complications and inferior survival. We conducted a single-arm study to test the feasibility and biological activity of a high-fiber, plant-based, whole-food meal delivery intervention during the peri-transplant period. Adults with multiple myeloma (n = 22) received fully prepared, plant-based meals for 5 weeks spanning conditioning, neutropenia, and early recovery, with the goal of supporting consumption of nutrient-dense, high-fiber foods despite transplant-related symptoms that often limit oral intake. The primary endpoints were feasibility and tolerability, defined by successful enrollment, adherence to study procedures, and patient-reported intake of study meals; diet was quantified using prospective food diaries and 24-hour dietary recall surveys. Secondary endpoints included changes in gut microbiome composition and function assessed by shotgun metagenomic sequencing and stool short-chain fatty acid (SCFA) measurements.

The intervention was feasible and generally well tolerated, with all participants consuming at least some proportion of delivered meals and with adherence sufficient to support planned dietary and correlative analyses. Greater intake of study meals was associated with more pronounced shifts in gut microbial communities, including enrichment of SCFA-producing taxa and compositional changes consistent with a fiber-responsive microbiome. Stool SCFA concentrations increased from baseline to the end of the intervention, suggesting a functional impact of the dietary strategy on microbial metabolite production during the peri-transplant period. These findings demonstrate that a plant-based meal delivery intervention is implementable during auto-HCT and suggest dose-dependent modulation of the gut microbiome and its metabolic output. Larger randomized trials are warranted to determine whether microbiome-targeted nutrition can reduce transplant-related toxicities, enhance immune recovery, and improve disease control in multiple myeloma. The trial is registered at ClinicalTrials.gov (NCT06559709).

## Introduction

Patients undergoing autologous hematopoietic cell transplantation (auto-HCT) for multiple myeloma face significant risks, including impaired immune recovery, infections, and eventually, almost inevitable disease progression. Emerging evidence suggests that dietary intake can influence the gut microbiome, which, in turn, affects immune function.^1^

The intestinal microbiome is markedly disrupted during cancer treatment and transplantation, and several clinical observational studies have demonstrated that the severity of this disruption may be a biomarker for outcome. In a large two-center observational analysis (n = 534), we demonstrated that auto-HCT patients arrive for transplantation with fecal microbial diversity that is lower than that of healthy volunteers and that patients subsequently incur further microbiome disruption during the neutropenic period following transplantation.^2^ Specifically, we observed that patients with greater microbiome damage, quantified by below-median fecal diversity during the peri-neutrophil-engraftment period, had higher rates of disease progression following transplantation and inferior overall survival compared with patients with preserved microbial diversity. Three other single-center studies reported similar findings.^3–5^

Prior work also suggests that plant-based eating patterns may benefit people across the myeloma spectrum. Large cohort analyses have found markedly lower multiple myeloma incidence among pescatarians, vegetarians, and vegans compared with meat-eaters (70–80% relative risk reduction, though based on few cases), suggesting a potential protective effect from plant-rich diets even before diagnosis.^6^ A recent trial in patients with myeloma-precursor conditions (monoclonal gammopathy of uncertain significance (MGUS) or smoldering myeloma) tested a high-fiber, plant-based dietary intervention in overweight or obese individuals for 24 weeks.^7^ Provision of plant-based meals paired with coaching was feasible and improved quality of life and multiple additional parameters, including body mass index, insulin resistance, and intestinal microbiome diversity and composition. Disease trajectories were stable or improved throughout the study period, and complementary experiments using the Vk*MYC mouse model of myeloma demonstrated that a high-fiber diet delayed disease progression via microbiome remodeling, increased short-chain fatty acid production (SCFA, e.g., acetate, butyrate, and propionate), and enhancement of antitumor immunity, supporting the hypothesis that high-fiber plant-based nutrition may be a viable strategy to slow disease progression in myeloma.

Mechanistically, plant-based diets contain fermentable fiber, which is a critical substrate for SCFA production by gut microbes. SCFAs have immunomodulatory capacity, and SCFA-producing taxa appear depleted in active and poor-prognosis myeloma, providing a plausible biological link. Conversely, western dietary patterns, which tend to contain a large proportion of processed meats, high-fat dairy products, and refined grains, have been shown to be pro-inflammatory and are associated with higher myeloma risk, reinforcing the rationale for emphasizing minimally processed plant-based foods.^8^ Furthermore, plant-based diets which contain a large proportion of high-fiber foods such as fruits, vegetables, whole grains, legumes, nuts and seeds typically have a higher nutrient density than western diets, and nutrient density has been demonstrated to lower the risk of cancer and all-cause mortality.^9^ Overall, while causality is unproven and data remain limited, encouraging patients to adopt a nutrient-dense, high-fiber, plant-based diet as a strategy for microbiome optimization and long-term favorable outcomes is supported by observational and early interventional findings in patients with myeloma and its precursor states.

Despite strong observational links between microbial health and transplant outcomes, no microbiome-directed dietary interventions are currently integrated into routine auto-HCT care. The central hypothesis of this work is that increasing intake of diverse, fiber-rich, plant-based wholefoods during the peri-auto-HCT period will optimize microbial health, mitigate the most common transplant-related toxicities (including infections), and favorably influence immune recovery and longer-term outcomes. To inform the design of a definitive randomized trial, we conducted a pilot study to deliver plant-based whole-food meals to patients undergoing auto-HCT for myeloma, given profound treatment-related disruption of oral intake and frequent gastrointestinal symptoms. This pilot study demonstrated that the intervention was feasible and well-tolerated by this patient population. Furthermore, we observed dose-dependent associations between consumption of study meals and changes in the microbiome, along with increased stool SCFA concentrations at the end of the intervention period compared with baseline, supporting further evaluation in larger, randomized studies.

## Methods

### Recruitment and Enrollment

The trial protocol was approved by the Fred Hutchinson Cancer Center IRB (FH/IRB0020501) and registered on ClinicalTrials.gov (NCT06559709). Recruitment and screening for eligible participants occurred from October 2024 to July 2025. Recruitment flowchart is shown in **Figure S1**. Inclusion criteria are described in the attached protocol. The majority who did not meet the inclusion criteria had medical comorbidities that necessitated a planned inpatient transplant, thus precluding home-delivered meals. Outpatient auto-HCT is the usual practice at our center. All interested prospective patients who met with the study team for consent visit agreed to participate, although 4 patients subsequently became ineligible (2 did not proceed to auto-HCT, 2 developed medical complications that necessitated inpatient admission for the duration of treatment). All participants provided written informed consent for their participation in the study, including collection, storage, and research use of their biospecimens and associated clinical data, in accordance with the Declaration of Helsinki. One participant declined to contribute sequencing data to public databases.

Additional control samples were obtained from participants enrolled in an institutional biobanking protocol approved by the Fred Hutchinson Cancer Center IRB (FH/IRB000956). All participants provided written informed consent for the collection, storage, and research use of their biospecimens and associated clinical data.

### Intervention

From day −7 to +28 (range: day −11 to +30), participants received three meals per day plus snacks, delivered twice weekly by Thistle, a commercial meal-delivery service. All meals delivered to participants were completely plant-based (with no meat, eggs, or dairy) and included breakfast, lunch, dinner, and a daily snack. The menu changed weekly, and the study dietitian chose each participant’s meals to adhere to the Fred Hutchinson Cancer Center Food-Safety Guidelines for Immunocompromised Patients, which are largely based on the US Food and Drug Administration recommendations for safe food handling and preparation. Per the food safety guidelines, pre-packaged salads, pre-cut raw fruits and vegetables, and raw nuts and seeds were not permitted. Breakfast options included: tofu scramble, oatmeal, and high-protein muffins. Lunches were one of four rotating soups, and dinners consisted of a plant-based protein source (e.g., tofu) with vegetables, legumes, and whole grains and pasta. Meals were delivered twice per week and required refrigeration, and minimal preparation (e.g., microwaving). Study participation did not mandate avoidance of meat, eggs, or dairy, nor did it preclude nutrition support (e.g., enteral or parenteral supplementation) if clinically indicated. Medical recommendations for nutrition support followed our usual care and were not altered for the study participants. Furthermore, all study participants were regularly assessed by an oncology-trained registered dietitian, in accordance with usual care at our center.

### Surveys, sample, and data collection

Dietary intake (of delivered meals as well as any other foods consumed) was monitored using a single 24-hour recall survey at baseline (prior to commencement of the food deliveries), nadir (d10 +/-5), and study end (d28 +/− 5), and via self-reported multi-day food records. Recalls were completed by the Fred Hutchinson Cancer Center Nutrition Assessment Shared Resource (NASR). Patients were also asked to complete multi-day food records at least three times per week throughout the study period, which were used to define consumption of the delivered meals. Participants met weekly with the study dietitian, during which dietary intake and tolerability were qualitatively assessed. Stool samples were requested weekly for exploratory analyses (6 in total throughout the study period). Standardized collection kits were provided, and patients returned the stool samples during clinic visits. Samples were processed within 24 hours and stored frozen at −80 degrees Celsius.

### Assessments and Endpoints

Primary endpoints were feasibility (enrollment, sample collection, and survey completion) and tolerability (meal consumption and qualitative patient feedback). We specified that we would deem the study feasible if we successfully recruited participants from our patient population and if 50% or more of the surveys and specimens were returned. To assess the tolerability of the intervention, the number of meals consumed, end-of-study survey data, and qualitative feedback from the study participants were evaluated.

Secondary endpoints were all exploratory. Nutritional exploratory endpoints included: 1) Adequacy of dietary intake, defined using accepted clinical nutrition guidelines (European Society for Clinical Nutrition (ESPEN))^10^ for calorie (30 kcal/kg/day) and protein (1.5g/kg/day) recommended minimums for this patient group. 24-hour dietary recall survey data were analyzed using the Nutrition Data System for Research (NDSR) software version 2024, developed by the Nutrition Coordinating Center, University of Minnesota, Minneapolis, MN. 2) Diet quality was assessed using the Healthy Eating Index–2020 (HEI-2020), modeled as a function of time and the intake of delivered meals. The HEI-2020 is a validated measure of overall dietary quality, and scores were generated by the NDSR software based on participants’ 24-hour dietary recalls. Microbiome-related exploratory endpoints included microbiome diversity and composition, abundance of metabolic pathway genes, and SCFA concentrations in stool. These were evaluated using shotgun metagenomic sequencing and metabolomics. Clinical data to support the analysis of exploratory endpoints were prospectively collected. Patient-reported outcomes were assessed using the EORTC QLQ-MY20 and QLQ-C30 at baseline and day 28.^11,12^

### Please see Supplementary Methods for Metagenomic sequencing, Metabolomics, Statistics, Software, and Data Sharing

## Results

### The study intervention was feasible and well-tolerated

Twenty-two patients were recruited over a 10-month period and commenced the study (**Table 1**). Greater than 50% of all planned biospecimen samples and patient surveys were successfully collected, indicating that the study procedures were feasible as defined in our study protocol. There were no withdrawals attributed to study-related procedures, and no adverse effects of the meal deliveries. The 5 patients who withdrew did so for logistic reasons related to hospital admission.

**Table 1.** Patient characteristics. Demographics of the study population.

| Table 1. Demographics of the study population. |  |  |
| --- | --- | --- |
|  | Age (median, years, range) | 58.5 (42-72) |
| **Sex** |  | n (%) |
|  | Female | 8 (36%) |
|  | Male | 14 (64%) |
| **Race** |  |  |
|  | Asian | 1 (4.5%) |
|  | Black or African American | 3 (14%) |
|  | White | 17 (77%) |
|  | Native Hawaiian or Pacific Islander | 1 (4.5%) |
| **Ethnicity** |  |  |
|  | Hispanic or Latino | 0 (0%) |
|  | Non-Hispanic or Latino | 22 (100%) |
| **R-ISS at diagnosis\*** |  |  |
|  | Stage 1 | 4 (20%) |
|  | Stage 2 | 13 (65%) |
|  | Stage 3 | 3 (13%) |
| **Disease stage at transplant** |  |  |
|  | CR | 1 (5%) |
|  | PR | 3 (13%) |
|  | VGPR | 18 (82%) |
| **ECOG** |  |  |
|  | 0 | 8 (36%) |
|  | 1 | 13 (59%) |
|  | 2 | 1 (5%) |

\*data missing to calculate R-ISS stage for 2 participants, so n=20 was used to calculate values.

Of the 22 patients who commenced the study, 17 received all 5 weeks of intended food deliveries (median days on study: 35; range: 7-35). An average of 112 meals per participant (range: 88 – 135) were delivered during the study period. Sixteen of these 17 patients returned the EOS survey. Notably, 13 of these patients required hospital admission for symptom management and did not consume the delivered meals during their inpatient stay. These admissions occurred during the neutropenic nadir period, where regimen-related toxicities (anorexia (65%), diarrhea (88%), dysgeusia (71%), mucositis (59%), and nausea (82%)) limited intake. Three patients (18%) received parenteral nutrition during hospitalization.

The median number of delivered meals that participants reported in the multi-day food records was 29 (range: 2-99; **Fig 1A**). This consumption followed a bimodal pattern, with the highest consumption during the pre-auto-HCT period and recommencement following neutrophil recovery in many patients (approximately half of the participants recommenced eating the meals). The expected peak of regimen-related toxicities coincided with a reduction in whole-food meal consumption and total calorie intake. Patients reported consuming a primarily liquid diet of oral nutrition supplements and soups (non-study foods) during this time window. Thus, despite variability in consumption across the different phases of auto-HCT, we deemed that the intervention was well-tolerated overall.

**Figure 1:**
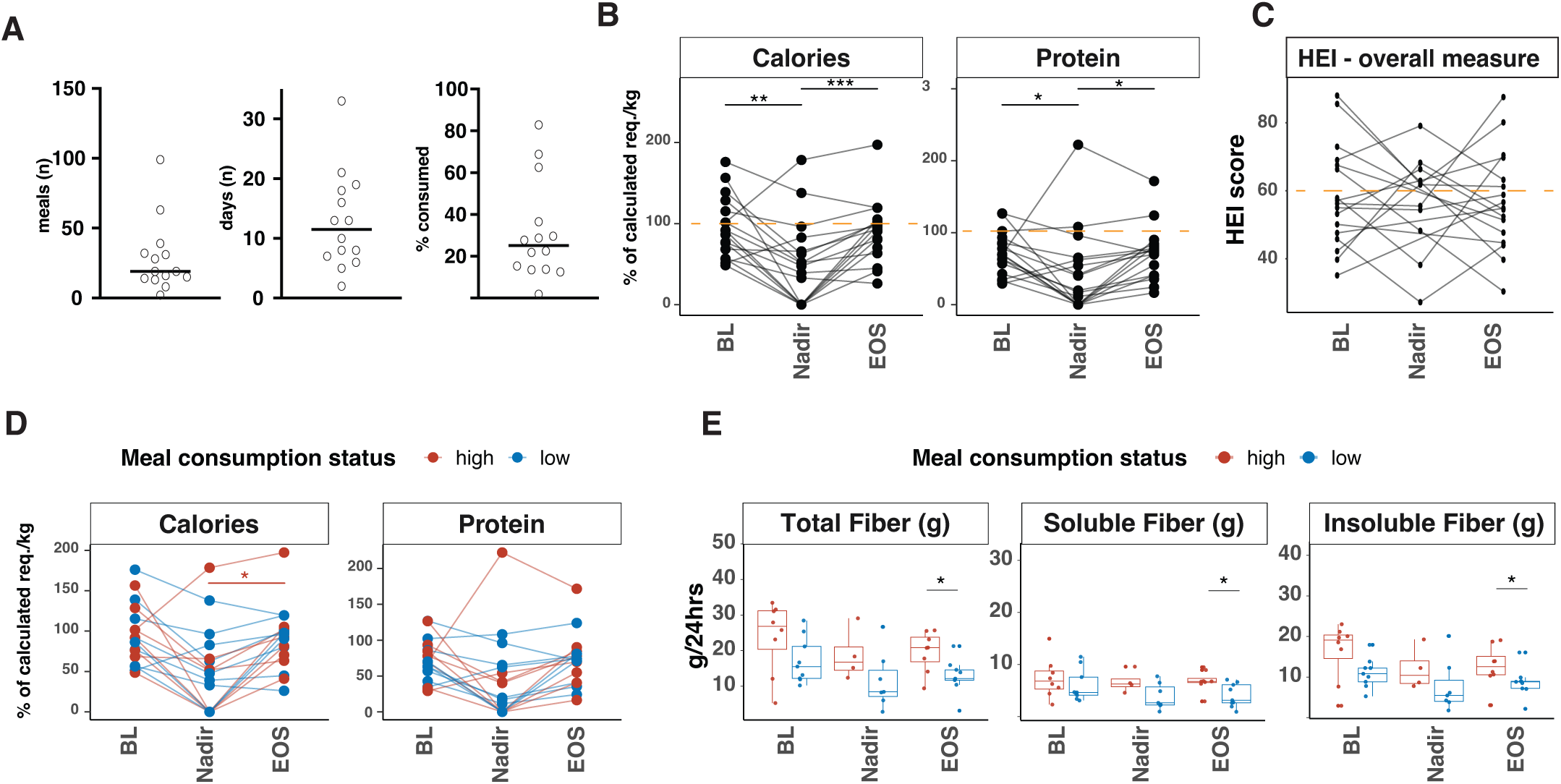
Fiber is increased during the study period in patients who are high consumers of plant-based meals. **A)** Summary of meal consumption: by total number of meals, days consumed, and proportion of total theoretical maximum, for 14 patients of the 17 who remained on study who returned food records. **B)** Fraction of recommended intake of calories and protein at baseline (BL), nadir (d10) and at the end of study (d28), as defined by 24-hour recall surveys. Dashed line highlights the 100% point. **C)** HEI index at BL, nadir and EOS, calculated from 24-hour recall surveys. Dashed line highlights the cut-point for the “very poor” range (<60). **D)** Fraction of recommended intake as a function of time and consumption status**. E)** Fiber intake status in high and low consumers over time. Statistical significance is indicated as P < 0.05 (*), P < 0.01 (**), and P < 0.001 (***).

### Dietary intake decreases during neutropenic nadir and calories can be increased during the post-nadir recovery period with high consumption of plant-based meals

Recommended nutrient needs for each patient were compared to nutrient intake collected from 24-hour dietary recall data at each timepoint (Baseline (BL), nadir and end-of-study (EOS)). Calorie and protein intake declined as expected^13^ during the neutropenic nadir (**Fig 1B**; plots show fraction of calculated needs consumed, where a value of 1 reflects meeting 100% of calculated needs). Dietary quality was analyzed using the healthy eating index standardized tool (HEI-2020), which is a scoring system that measures how closely a person’s overall diet aligns with the Dietary Guidelines for Americans, providing an overall diet quality score from 0 to 100. The average score in the general population is in the ‘very poor’ range (<60), and our patient group was similar to the general population, with scores <60 that did not significantly change over the study period (**Fig 1C**).^14–16^

The variation in meal consumption among patients provided an opportunity to divide the study cohort into ‘high’ and ‘low’ meal consumers (above and below median number of meals consumed) and explore the association between meal consumption and nutritional parameters. Several grouping strategies yielded very similar groupings, re-classifying only 3 of 14 participants between high to low groups, including absolute meals consumed, the number of days on which the plant-based meals were eaten, and the proportion of total meals consumed (of maximum delivered). For three patients who did not return food records, qualitative records from weekly interviews with the study dietitian allowed us to classify them into the low-consumption group.

High consumers had a higher calorie intake relative to predicted needs at the end of the study (p = 0.023, intake defined using the 24-hour recall surveys) compared with their neutropenic nadir, and this was not seen in the low consumers (**Fig 1D**). The high and low consumers did not differ statistically in calorie or protein intake at baseline. HEI did not differ between the high and low consumption groups, either in overall score, or specific vegetable and fruit sub-components (**Fig S1**).

When specifically evaluating fiber, ‘high’ consumers of the plant-based meals had a significantly higher total fiber intake at the end of study time point relative to the low consumers (**Fig 1 E**). This was not limited to soluble or insoluble fiber subtypes, as both were significantly increased.

### Microbiome analysis reveals several differentially abundant taxa both over time in the whole cohort, and in association with degree of plant-based meal consumption

We first visualized microbial composition using stacked bar plots, annotated by time relative to transplant and ‘high’ vs. ‘low’ meal consumption status (**Fig 2A**). Of note, eight patients (47%) received cefepime during the peri-transplant period. The average duration of cefepime exposure was 1.65 days (SD: 2.18 days). One patient received piperacillin-tazobactam for one day only. No patients received carbapenem antibiotics during the study period.

**Figure 2:**
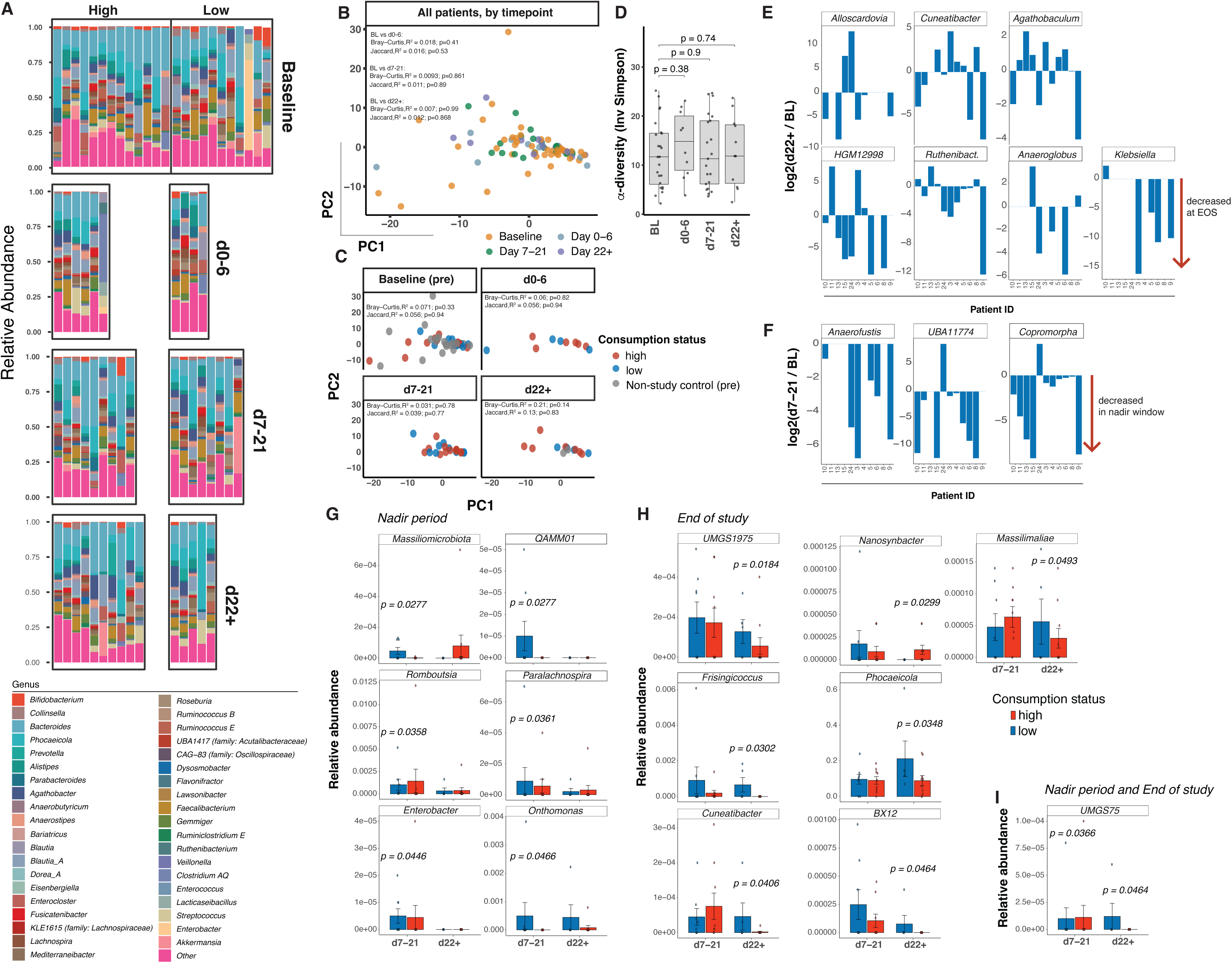
Plant-based meal consumption alters the intestinal microbiome during the post-auto-HCT recovery period. **A)** Bar plots showing overall microbiome composition by group and time point. Each bar represents a single stool sample. Where multiple samples were provided by the same patient for a given time window, all are included. **B)** PCoA plots colored by consumption status and divided by time point. **C)** PCoA plots demonstrating overall microbial composition at the pre-transplant time point, d0-6, d7-21, or d22+. High consumers are shown in red, low in blue. 15 unique additional control patients who were pre-auto-HCT but not study participants are shown in grey in the pre-transplant plot. All samples shown visually, but repeated measures are considered in PERMANOVA testing as annotated on the plots; genus-level data shown. **D)** Alpha-diversity (inverse Simpson index). Baseline n = 24; d0-6 n = 10; d7-21 n = 21; d22+ n = 11. **E)** Log2 fold-change analysis highlighting statistically significant changes between end-of-study (d22+) and baseline, all taxa shown p<0.05. **F)** Log2 fold-change analysis of changes occurring between nadir and baseline; all taxa shown p<0.05. **G-I)** specific taxa altered when both consumption status and time point are considered. Exploratory analysis, taxa with uncorrected p-values of <0.05 are shown. **H)** End-of-study, the single statistically different taxa at both time points.

To reduce dimensionality and identify broad trends, we next visualized microbiota composition using principal co-ordinate analysis with the Bray-Curtis dissimilarity (PCoA; **Fig 2B**). We observed relatively tight clustering over time, with some drift seen by d22+ (**Fig 2B**); meal consumption status explained 6.8% of the variance in gut microbiome community composition by the end of the study (PERMANOVA, Bray-Curtis, R^2^=0.068, p=0.08). When we considered each time comparison alone (agnostic of exposure status), there were no trends in either Bray-Curtis or Jaccard analyses (**Fig 2B**). Furthermore, we did not observe differences between the high and low meal consumers during each time window, likely due to limited sample sizes (PCA shown in **Fig 2C**). Furthermore, the inclusion of 15 additional control samples, obtained from non-study patients undergoing auto-HCT for myeloma who participated in a biobanking protocol, suggested that the study cohort did not differ from the general pre-auto-HCT pool at baseline (**Fig 2C**).

We did not observe the decrease alpha-diversity that has been shown in other studies during the peri-engraftment (d7-21) period (**Fig 2D**). This may be explained by small numbers in this cohort, however it is also possible that the maintenance of baseline microbial features is explained by the dietary intervention administered, even in those who were not high consumers (**Fig 2D**).

To further understand whether any compositional differences developed during the study, we inspected fold-changes in relative abundance of taxa between baseline and end-of-study (independently of intake status), which highlighted several genera that are significantly different (paired analysis, per-patient data shown in **Fig 2E**, all p<0.05). These genera were largely anaerobic commensals in phylum Firmicutes: *Agathobaculum* (family Butyricicoccaceae), *Cuneatibacter* (family Lachnospiraceae), HGM12998 (an uncharacterized Oscillospiraceae genus with presumed roles in complex carbohydrate metabolism and SCFA production and not considered to be a pathobiont),^17^ *Anaeroglobus* (family Megasphaeraceae) and *Ruthenibacterium* (family Ruminococcaceae), and the Actinobacteriora phylum member *Alloscardovia* (family Bifidobacteriaceae). Alloscardovia is closely related to classical Bifidobacteria and is typically an inhabitant of the gastrointestinal and genitourinary tracts; it participates in carbohydrate fermentation and is usually considered a low-virulence commensal, although occasional opportunistic infections have been reported, particularly in medically complex hosts.^18,19^ *Klebsiella* (family Enterobacteriaceae) represents a facultatively anaerobic genus within phylum Pseudomonadata (previously Proteobacteria) that can expand during dysbiosis and is well recognized as an opportunistic pathogen causing bloodstream infections in immunocompromised patients.^20^ We observed a statistically significant reduction in Klebsiella abundance at the end of study compared with baseline samples, suggesting a potential for plant-based dietary approaches in reducing the burden of pathobionts and thus subsequent infections.

We further analyzed shifts between baseline and the nadir period (d7-21), as this window has been most predictive of outcome in previous studies of HCT, and here there were 3 taxa that were significantly decreased compared with baseline: *Copromorpha* (family Anaerovoracaceae), *Anaerofustis* (family Anaerofustaceae)^21^ and UBA11774 (an as-yet uncultivated member of the Lachnospiraceae family; **Fig 2F**). These three taxa are all obligate anaerobic Firmicutes, and their loss during the nadir period post auto-HCT likely reflects the dysbiosis that is not fully reflected in alpha or beta-diversity metrics in this small cohort.

We next explored fold-change analyses at the nadir time window (d7-21) and at the end of study (d22+), focusing on identifying differences between high and low consumers of the delivered meals, considering the interaction between time and consumption status in our statistical analysis. These data highlight that high consumption of plant-based foods may shift the microbiome in a favorable direction, including by increasing the abundance of healthy commensal bacteria (relative abundance plots, **Fig 2G-I**, with annotated p values derived from log2 fold-change analysis). Several of the identified taxa represent uncultured bacteria that have been isolated from the human gut but have not yet been assigned formal Latin names. UMGS1975, of family QAND01 within the order Christensenellales, phylum Firmicutes A (also called Bacillota A) was the most significantly different between the high and low consumers at the end of study and was elevated in low consumers.

### Microbial pathways linked with SCFA production are differentially abundant in the high consumers of plant-based diets

We next explored the functional consequences of these changes in microbial taxa. We identified metabolic pathways that were differentially present in the high and low consumers of delivered meals at both the neutropenic nadir period and at the end of the study (**Fig 3A-B**). Notably, the most significantly increased pathway in the high consumers at both time points was aminobutanoate degradation (PWY5022), that directly results in SCFA production (e.g., butyrate), and initiation of fatty acid biosynthesis in mitochondria (PWY66-429) (**Fig 3C**). Other differentially abundant pathways are listed in **Table S1**.

**Figure 3:**
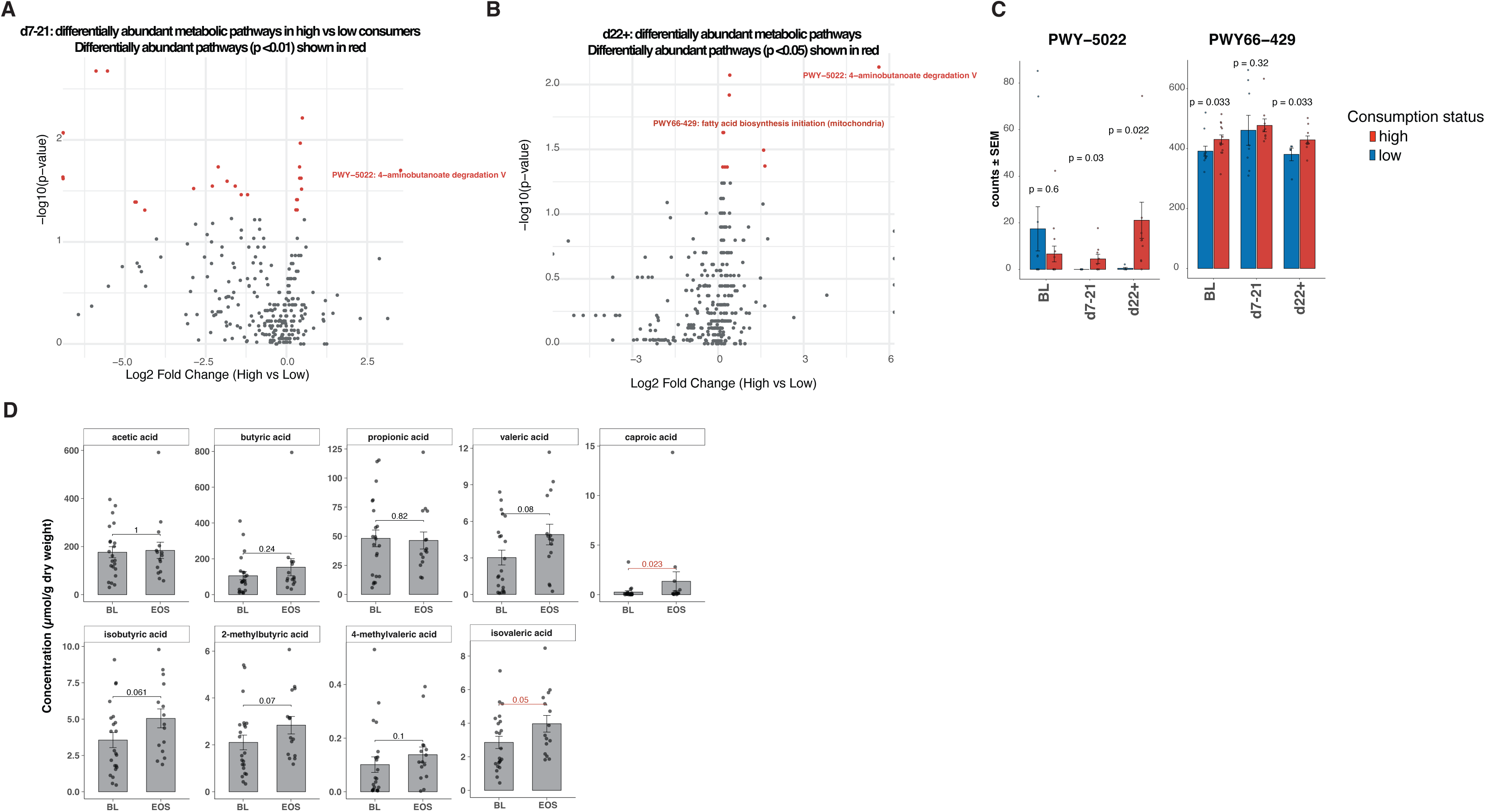
Stool SCFA are increased at the end of the study period compared with baseline samples A, **B)** Volcano plots showing differentially abundant metabolic pathway genes (log2 fold-change) in samples collected between **A)** d7 and 21 post-auto-HCT and **B)** after d22, considering meal consumption status. **C)** Two specific pathways related to SCFA production, during the pre-transplant, nadir, and end-of-study time windows. P-values comparing low and high consumers at each time point shown. 22 samples included at BL timepoint, 17 during neutropenic nadir, and 15 at EOS. **D)** SCFA were measured in stool samples at baseline and at end of study. All 22 patients gave a baseline stool sample, and 15 completed an end-of-study sample; last available was used where required. Wilcoxon non-parametric tests were performed.

We next measured SCFA in stool samples from baseline and at the end of study. We observed an increase in SCFA within the patient cohort as a whole, with caproic acid and isovaleric acid reaching statistical significance (p = 0.023 and 0.05 respectively), and valeric acid, isobutyric acid and 2-methylbutyric acid reaching near-significant values (p = 0.08, 0.06, and 0.07, **Fig 3D**). Expectedly, given the small patient numbers, we did not observe differences in the measured metabolites when we compared the low and high consumption groups.

### Role functioning improves throughout the study period, while other aspects of QoL remain unchanged or worsen

Patient-reported outcomes were also measured using the myeloma-specific MY20 and general cancer-related C30 tools, and confirmed that some aspects of quality-of-life improved throughout the study period. The MY20 (EORTC QLQ-MY20) is a 20-item, myeloma-specific quality-of-life questionnaire used alongside the EORTC QLQ-C30 to assess domains such as disease symptoms, treatment side effects, future perspective, and body image in patients with multiple myeloma, with each domain scored from 0 to 100 so that higher scores on symptom and side-effect scales indicate worse symptom burden while higher scores on functioning-oriented scales (e.g., future perspectives, role functioning) indicate better quality of life. Analysis of the MY20 demonstrated an increase in disease symptom scores but an improvement in scores related to side effects of treatment and body image. Scores for future perspectives were unchanged throughout the study period. Analysis of the C30 survey results demonstrated that scores for global health status/QoL declined between the baseline assessment and the end-of-study (day 28 post-auto-HCT), but there was a modest improvement in physical and social functioning, a substantial improvement in role functioning, and no changes in emotional and cognitive functioning. The role functioning metric captures limitations in work, household tasks, and other usual activities due to physical or emotional barriers. Specific scores are summarized in **Table S2**. Overall, the patient-reported outcomes provide valuable insights into the patient experience of auto-HCT, and we will build on these findings in future studies.

## Discussion

This pilot study demonstrates that a home-delivered, plant-based meal intervention during auto-HCT is feasible and generally well tolerated, though strategies to improve adherence will be important for future studies in this patient group. Higher meal consumption was associated with improved calorie and fiber intake, favorable shifts in select commensal gut taxa, and enrichment of microbial pathways linked to SCFA production. Together, these findings support the potential of targeted, plant-based nutritional strategies to modulate the gut microbiome and metabolic outputs during auto-HCT, warranting confirmation in larger, controlled studies.

The intervention met its primary feasibility endpoint, demonstrating that patients undergoing auto-HCT can participate in a plant-based whole-foods meal program during an intensive treatment period. While GI symptoms did limit intake during the nadir period, most patients recovered, and oral intake increased with count recovery. Future studies will include additional meal options for patients that are easier to swallow, such as smoothies and simple soups during the neutropenic phase of the transplant.

This is the first study to longitudinally explore HEI during the peri-transplant period. Our study population had an HEI score in the ‘very poor’ range throughout the study period, and while this is in line with national US averages, it is likely that the pre-transplant diet quality reported in our patients is influenced by several factors unique to this patient population. Pre-transplant auto-HCT patients have typically just completed induction-phase therapy and are experiencing residual GI toxicities, and many reside away from home to be treated at our specialized cancer center and thus have limited access to cooking facilities. The low HEI scores highlight the importance of dietary intervention strategies in this patient population, as well as the challenge in improving dietary quality and diversity from a low baseline.

While this study was small, and microbial and metabolic studies were exploratory, we still observed several changes within the study participants both over time and as a function of meal consumption. Consumption of the delivered plant-based meals was associated with measurable shifts in gut microbiome composition, with the largest changes observed among participants with the highest intake of plant-based study meals. Critically, end-of-study fecal samples demonstrated an increase in SCFA within the intestinal tract, which suggests dietary intake can modulate microbial community function, even in the setting of post-HCT tissue damage. SCFAs are known to support epithelial barrier integrity and modulate immune function, providing a mechanistic rationale for targeting the microbiome with diet in myeloma.

Plant-based dietary patterns typically emphasize vegetables, whole grains, fruits and legumes, nuts and seeds and make these foods the primary focus of each meal (typically >75%). However, individual interpretations of this term vary, making strict definitions difficult. The delivered foods were strictly of plant origin. However, we deliberately did not require that patients exclude animal foods from their diets to participate in this study, as this would not have been deemed acceptable by our patient population.

There were several limitations to our study. Firstly, while we have assessed that the intervention is tolerable overall, future studies will require strategies to improve intake during the neutropenic nadir, when gastrointestinal symptoms are at their maximum. These include: increasing the availability of more familiar ‘comfort foods,’ as well as soups, smoothies, and other nutritional supplements. For our predominantly outpatient auto-HCT approach, we also need to develop a strategy to continue meal support during inpatient hospitalization. Given the dose-dependent findings, we hypothesize that improving meal intake will lead to even greater nutritional and microbiome benefits.

Overall, we conclude that the provision of plant-based whole-food meals can alter the gut microbiome in a dose-responsive fashion and is associated with increased short-chain fatty acid (SCFA) production, supporting larger efficacy-focused trials. In future studies, we propose incorporating behavioral and implementation science to optimize adherence and to test whether functional microbiome changes mediate improved transplant and myeloma outcomes.

## Supporting information

Trial protocol

## Data Availability

All data produced in the present study are available upon reasonable request to the authors and sequencing and metabolomic data will be made publically available within the appropriate NIH databases at the time of formal publication.

## Supplementary Materials

**Figure S1:**
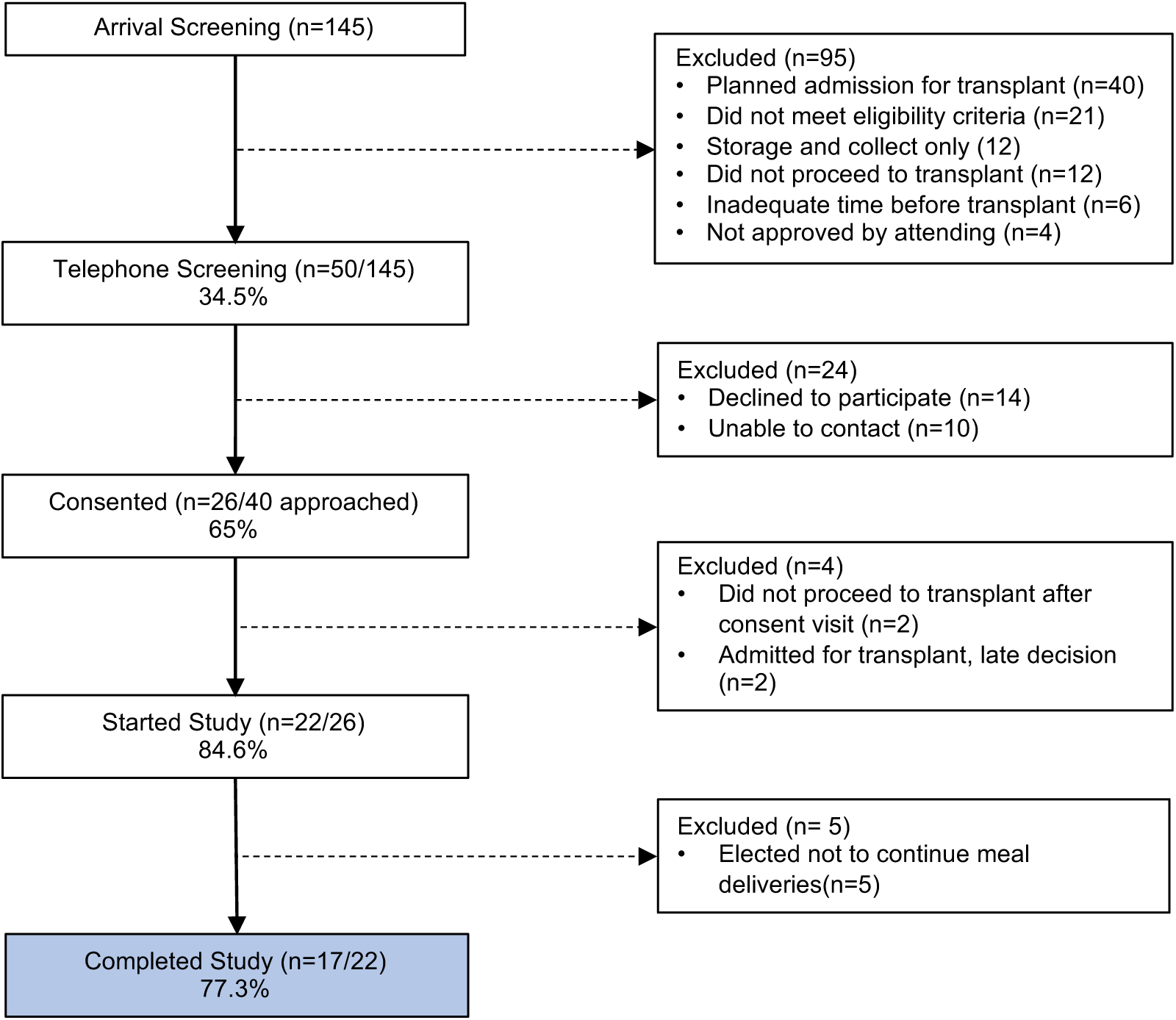
Accrual summary.

**Figure S2:**
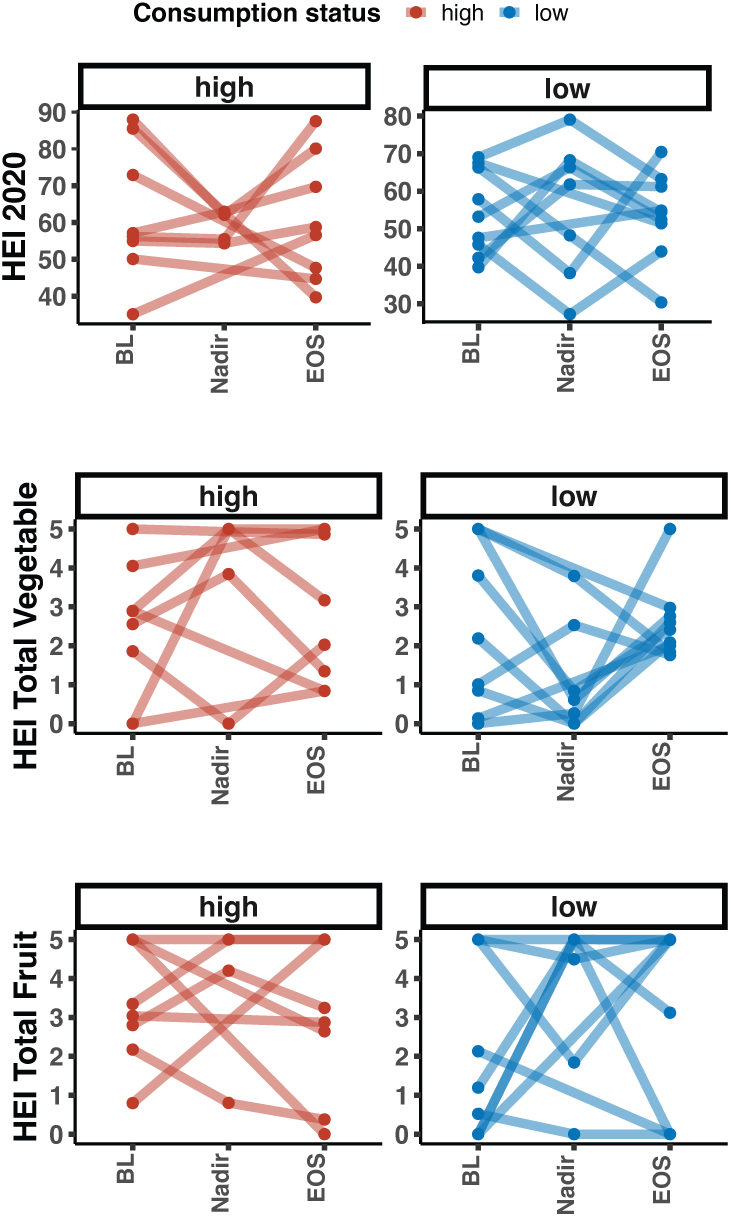
Sub-scores of the HEI. HEI index at BL, nadir and EOS, calculated from 24-hour recall surveys, subsetted by high (n = 7) and low consumption status (n = 10). Total HEI and the Vegetable and Fruit components shown.

**Table S1:**
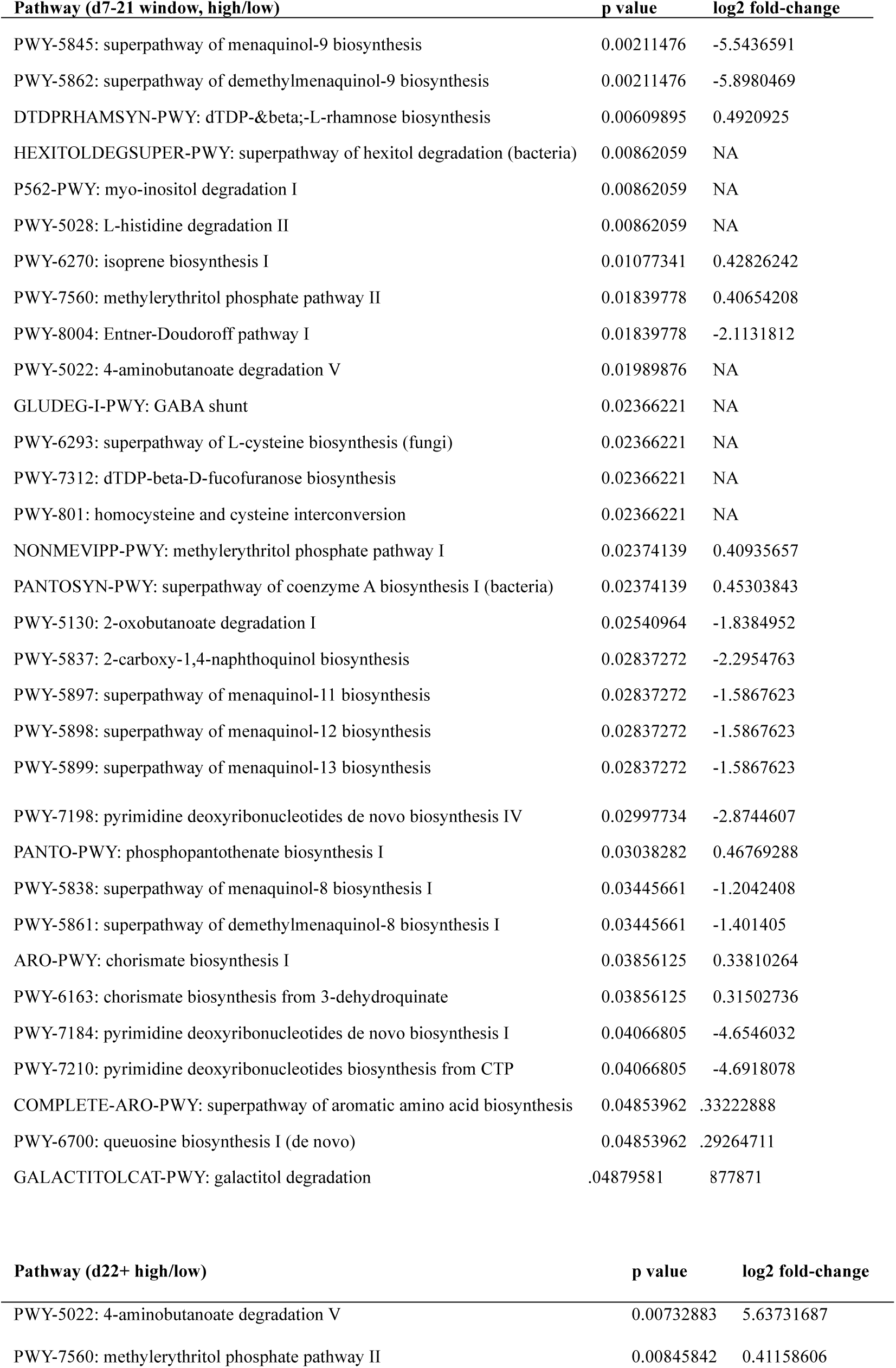
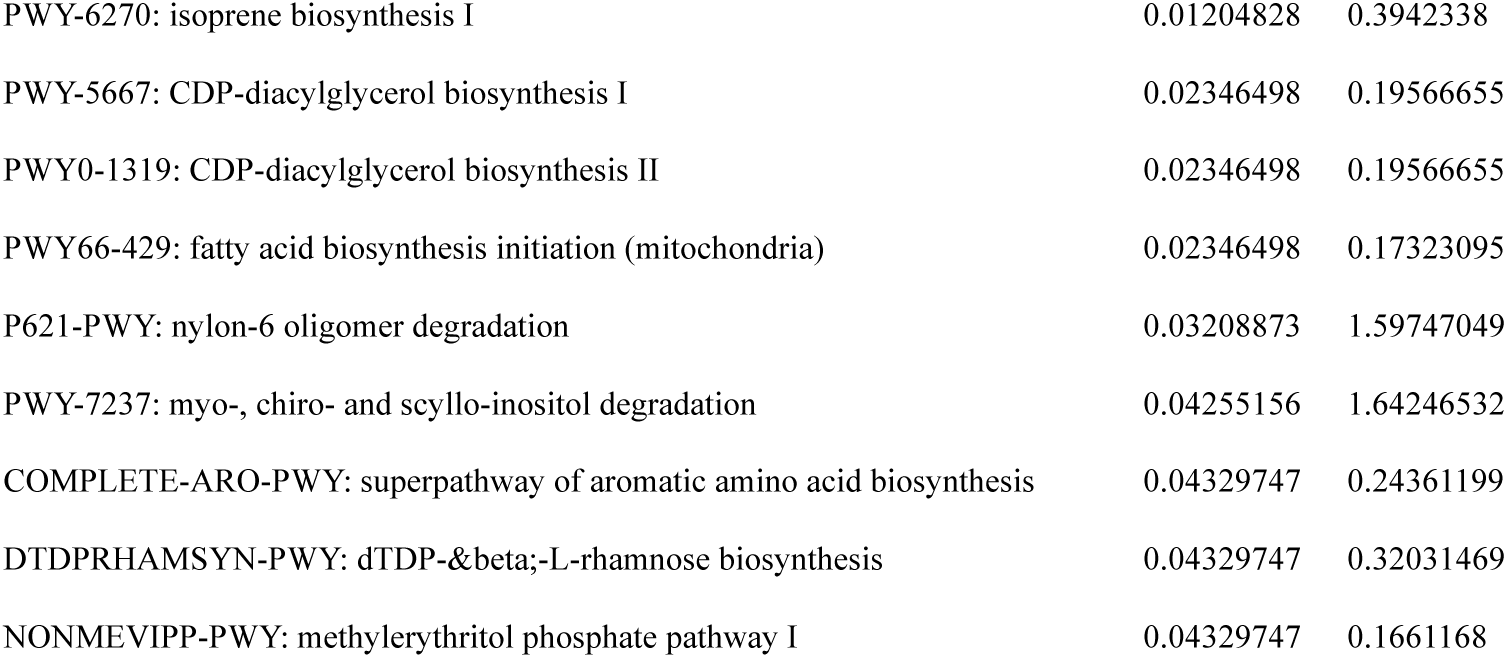
Differentially abundant metabolic pathways (Volcano plots shown in Figure 3)

**Table S2:**
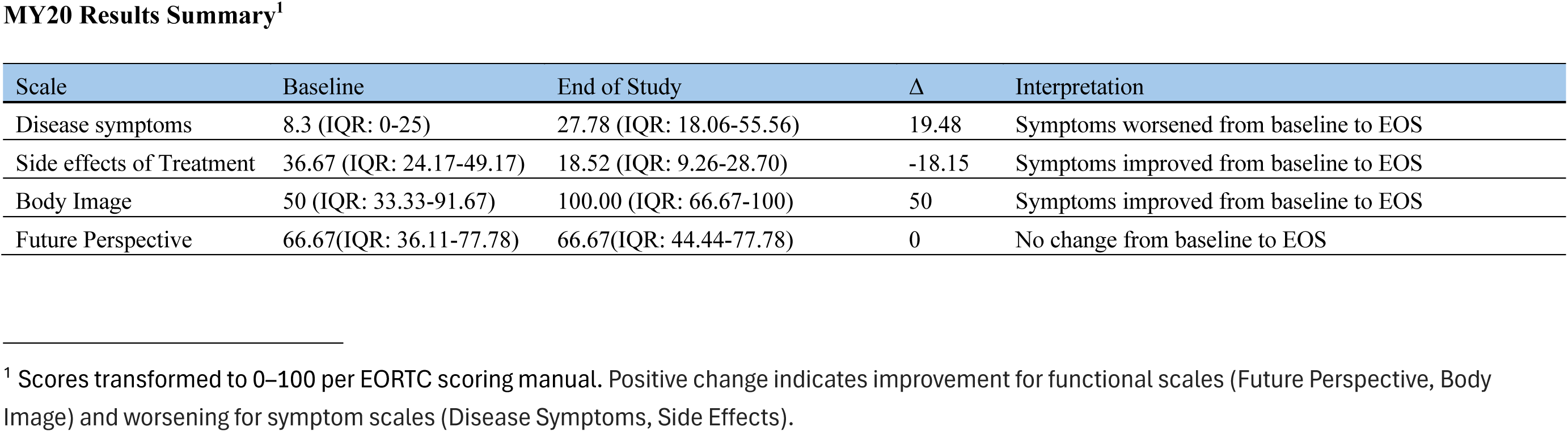
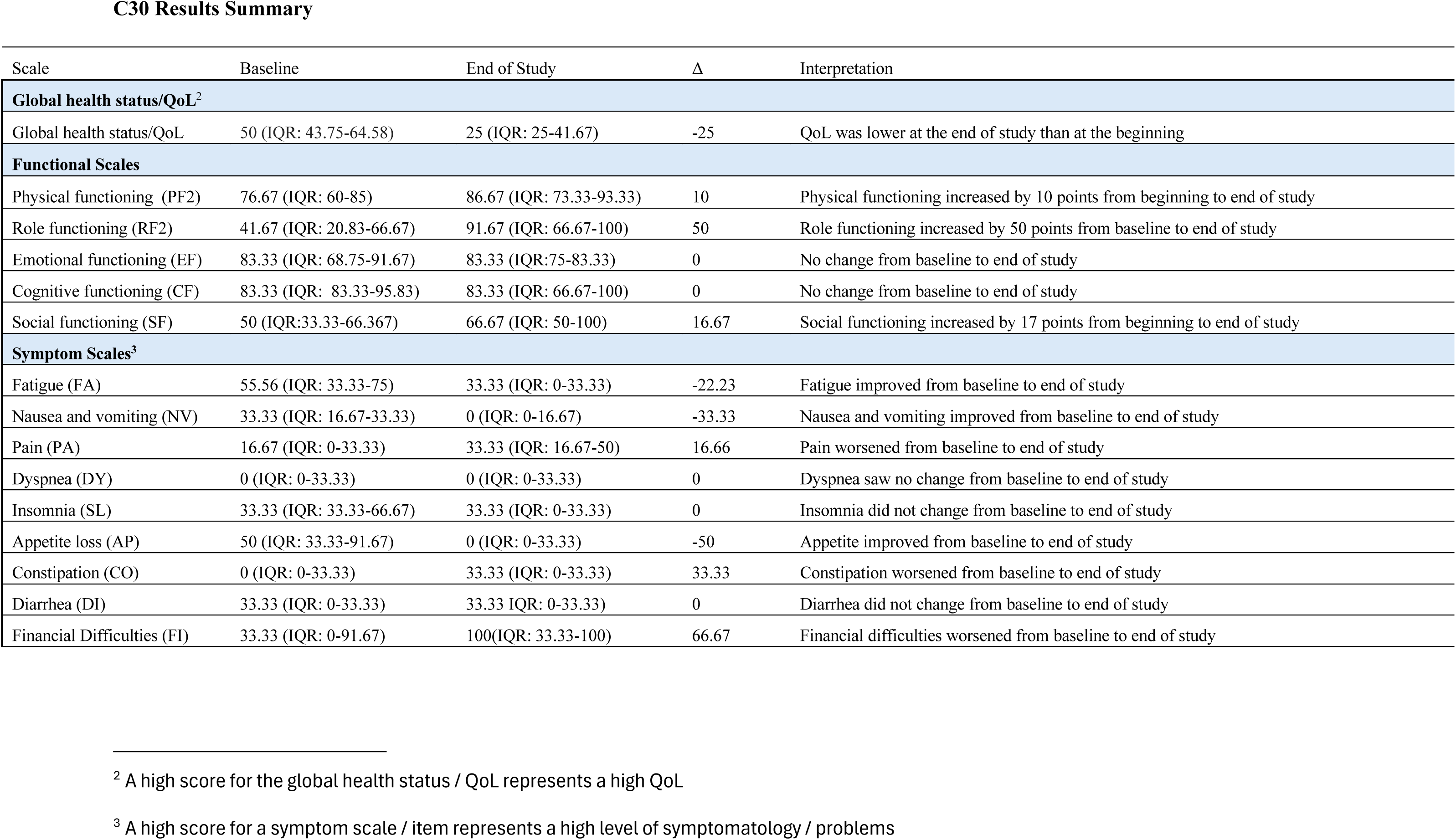
MY20 and C30 scores.

