## Supplementary material for "Plant-based whole-food diets are feasible during autologous stem cell transplantation and are associated with dose-dependent microbiome modulation: Results from a pilot clinical trial": Trial protocol

Fred Hutch IRB
Approved
8/7/2024

### Implementing a plant-based whole-foods meal delivery service for patients undergoing autologous hematopoietic cell transplantation for Multiple Myeloma: A pilot study

**Principal Investigator:**

Kate Markey, MBBS, MClinRes, PhD, FRACP
Assistant Professor, Fred Hutchinson Cancer Center, [redacted]

**Sub-Investigators:**

Stephanie Lee, MD, MPH
Professor, Fred Hutchinson Cancer Center, [redacted]

Leona Holmberg, MD, PhD
Professor, Fred Hutchinson Cancer Center, [redacted]

David Fredricks, MD
Professor, Fred Hutchinson Cancer Center, [redacted]

Kate Ueland, MS, RD, CSO
Lead study Research Dietitian, Fred Hutchinson Cancer Center

Kerry McMillen, MS, RD, CSO, FAND
Medical Nutrition Therapy Manager, Fred Hutchinson Cancer Center, [redacted]

Mary Rasmussen, MS, RD, CSO
Dietitian, Fred Hutch Cancer Center

Mariia Byelykh, MS, RD, CD, CNSC
Clinical Dietitian, Adult Blood & Marrow Transplant, Immunotherapy, and Oncology Service

Study statistical advisor: Ted Gooley

Fred Hutch IRB
Approved
8/7/2024

Fred Hutch IRB
Approved
8/7/2024

### 1 INTRODUCTION

#### 1.1 Background and Rationale

High-dose chemotherapy followed by autologous stem cell transplantation (ASCT) remains the standard-of-care consolidation treatment for multiple myeloma (MM).[1, 2] Monotherapy with high-dose melphalan is the standard conditioning regimen for MM.[2]

The process of ASCT commonly leads to the following effects for patients: chemotherapy-induced mucositis, malabsorption/diarrhea, changes in nutritional intake, and often a need for broad-spectrum antibiotics, most commonly as empiric therapy for febrile neutropenia. These changes and interventions, specifically antibiotic therapy and dietary perturbation, alter the intestinal microbiome, a complex community consisting of trillions of microbes that inhabit the mammalian gastrointestinal tract and regulate host physiology.[3, 4] In healthy individuals, diet intervention with fermented foods, for example, has been shown to improve microbiota diversity, reduces inflammation, and modulates immune responses.[5] In patients undergoing initial therapy for MM, a plant-based whole foods (PBWF) diet has been linked to an improvement in bacterial diversity within the intestinal tract, and a long-term PBWF diet is associated with improvements in QOL, fatigue scores, and insulin resistance. In an observational study published in 2020, Dr Shah and colleagues at Memorial Sloan Kettering Cancer Center reported that the intake of seafood and plants was positively correlated with the short-chain fatty acid butyrate in stool samples, as well as sustained MRD-negativity in myeloma patients receiving lenalidomide maintenance therapy.[6]

We recently showed that ASCT patients sustain severe microbiome injury during the course of transplantation and that low diversity of the fecal microbiota community predicts poor progression-free and overall survival.[7] Specifically, in a large two-center observational analysis (n = 534), we showed that ASCT patients arrive for transplantation with fecal microbial diversity that is lower than that of healthy volunteers and that patients subsequently incur further microbiome disruption during the neutropenic period following transplantation. This disruption is marked by low alpha-diversity (**Figure 1**) with domination of fecal communities by potentially pathogenic organisms, most notably members of the Streptococcus and Enterococcus genera. In concert with our analysis, which was the largest and first multicenter microbiome study in ASCT, three smaller single-center studies reported similar findings.[8-10] Critically, we observed that patients with greater microbiome damage, as assessed by below-median fecal diversity around the time of neutrophil engraftment, had higher rates of progression following transplantation and inferior overall survival (**Figure 2**).[7] These patterns of microbiota disruption are similar to changes during allogeneic hematopoietic-cell transplantation, where loss of diversity is associated with inferior outcomes.[11]

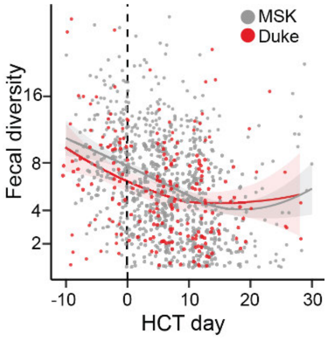

**Figure 1.** Fecal diversity declines in auto-HCT. 841 samples from 384 patients from two centers. Modified from Khan et al, Blood 2021.

Fred Hutch IRB
Approved
8/7/2024

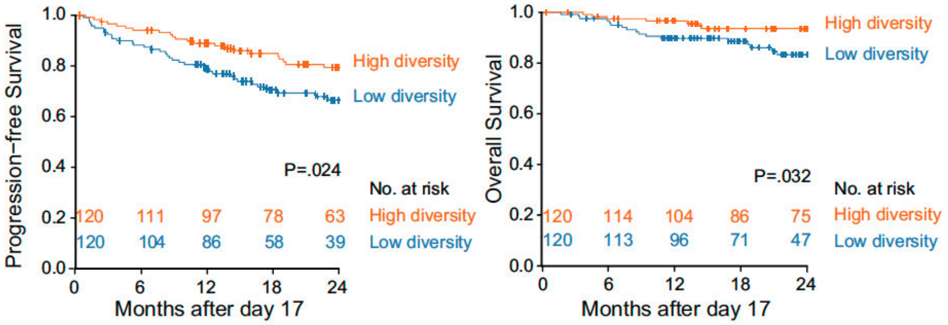

**Figure 2.** ASCT patients with higher diversity in fecal samples collected between HCT days 9 to 16 are at a reduced risk of progression or death. Modified from Khan et al, Blood 2021.

The immune system participates in MM and lymphoma disease control,[12-16] and impaired immunity occurs with disease evolution.[16-18] Furthermore, immune dysfunction, including T-cell exhaustion, after ASCT contributes to relapse,[19, 20] and interventions to prevent T-cell exhaustion may counter MM disease progression after ASCT.[20] Importantly, a diverse intestinal microbiota has beneficial immunomodulatory properties[21] that may mitigate relapse after ASCT by providing locally acting and circulating metabolites that support “T-cell health.” We previously reported that stool samples from MM patients on maintenance therapy without minimal residual disease had higher relative abundance of *Eubacterium hallii* than those with detectable disease and identified *Eubacterium limosum* and *Faecalibacterium prausnitzii* as candidate taxa implicated in freedom from MM progression.[22, 23] Support for a direct effect of the gut microbiota on MM progression was demonstrated in a murine study in which MM progression was modulated by manipulating the intestinal microbiome and promoting the expansion of IL-17-producing cells.[24] Also, syngeneic adoptive cell therapy depends in part on signals from the intestinal microbiota crossing an impaired gut barrier to augment anti-tumor immunity,[25] supporting a contribution of circulating metabolites to immune modulation.

Despite our observation that microbial health is linked with optimal outcome for transplant patients, there are no targeted interventions currently in clinical practice. Within the autologous stem cell transplant recipient population, there is also a profound lack of clinical trials, meaning that we will be offering a unique approach in this study.

We hypothesize that increasing intake of a diverse variety of plant-based whole foods, high in fiber and micronutrients, will improve microbial health during the early post-autograft period. We propose that this, in turn, will have beneficial effects on transplant-related toxicities (e.g. diarrhea) but also on immune function, and thus long term outcome.

### 2 OBJECTIVES

Our ultimate goal is to determine if a plant-based whole foods diet that is high in fiber and a variety of micronutrients can prevent microbiome damage during ASCT, which may, in turn, lead to other benefits. The first step in this process is to explore the feasibility, safety, and acceptability of this approach. We will do that by performing this **single-center, single-arm pilot study of 5 weeks of a plant-based whole foods diet during the peri-transplant period, starting the week before ASCT.** Patients will receive meal deliveries (breakfast, lunch, dinner, and snack options) from a commercial company (Thistle), whom we will partner with for the trial.

Feasibility measures that we seek to confirm:

- Determine the proportion of patients who consent to and enroll in the trial
- Establish the proportion of the delivered meals and snacks eaten throughout the course of the study
- Determine the acceptability of the plant-based whole foods diet and

Scientific correlative goals:

- Profile stool samples collected before and throughout transplant using shotgun metagenomic sequencing
- Measure concentrations of short-chain fatty acids in stool (e.g. butyrate) to determine if initiating and maintaining oral fiber intake (or via enteral nutrition, if clinically indicated) leads to increased production of microbe-generated metabolites known to support gut health
- Profile the recovering post-transplant immune system using multiparameter flow cytometry in order to link the microbial profile with immune cell phenotypes, especially of recovering T cells.

Additional exploratory analyses:

- Quantify the proportion of each meal eaten using food records (multi-day food record), and establish the frequency of patients recording their oral intake.
- Use a 24-hr recall analysis of all foods eaten at 3 defined time points (enrolment, during neutropenic nadir and at study end) to quantify the number of different foods eaten during each window.
- Explore the relationships between clinical correlative analyses of stool and blood with dietary intake and outcomes.
- Pilot the use of data analysis methods in preparation for a future randomized study.

This is a single-arm pilot study. We will compare our correlative findings (immune profiling, stool) with historical control data from our own center and from the published literature, including work previously published by the study PI. We will also use the outcome data (disease response at 30 days and 1 year) to guide the design of a randomized study in the future.

### 3 RESEARCH SUBJECT SELECTION

#### 3.1 Inclusion Criteria

*Subjects must meet all the following criteria at enrollment:*

- Able to provide written informed consent prior to initiation of any study procedures
- Planned first autologous stem cell transplantation for multiple myeloma

Fred Hutch IRB
Approved
8/7/2024

- Planned outpatient treatment for the duration of transplantation (if admitted, we will request that caregivers bring the meals/snacks to the hospital as they might with other food prepared at home).
- Access to a refrigerator
- Ability to reheat foods
- Able to consume an oral diet at enrolment
- Able to communicate clearly regarding aspects of the study: *e.g.* Give feedback on logistics and meals, in order to maximize the operational data we can gather in this pilot study
- At least 18 years of age

##### 3.2 Exclusion Criteria

*Subjects must not meet any of the following criteria at enrollment:*

- Major psychiatric diagnosis that impairs cognitive functioning or is not controlled at the time of the approach, as judged by the patient’s medical team
- Planned inpatient transplantation (we will accommodate meal delivery to UWMC for planned outpatients who get admitted, but we will restrict the pilot study to planned outpatients for logistic reasons).

#### 4 RESEARCH PARTICIPANT ENTRY

Prior to the implementation of this protocol, and any subsequent full version amendments, the protocol and the protocol consent form(s) will be approved, as appropriate, by the institutional review board (IRB). All potentially eligible patients scheduled to receive autologous HCT will be identified and screened via chart review. A member of the study team will approach the patient and provide a description of the study and the requirements for study participation. Patients will be given an opportunity to ask questions. If a patient agrees to participate, they will be consented by the Primary Investigator or a Sub-Investigator and the patient will sign and date the current, approved consent. The consent form will describe the purpose of the study, the procedures to be followed, and the risks and benefits of participation. A copy of the consent form will be given to the participant and consent will be documented in the participant’s record. If a patient declines participation, the reason for their decision will be elicited and noted so that we can better understand the feasibility of the study.

We seek to enroll and be able to evaluate 20 patients. For those who decline, we will document reasons for non-participation, and the approach to recruitment will be reevaluated for future patients and subsequent trials. We will replace participants who drop out of the study. Of note, all patients, whether enrolled in this study or not, meet with the clinical registered dietitian upon arrival to the transplant team as part of their clinical care. All patients will also be followed by a study-associated registered dietitian weekly while on the study.

#### 5 Study Design

Prior findings in the field beg the question: How can we intervene to improve intestinal microbial diversity in myeloma patients undergoing ASCT?

Fred Hutch IRB
Approved
8/7/2024

This is a feasibility study of delivering plant-based whole foods meals to patients during the peri-transplant period in order to encourage diverse food consumption of high-fiber, nutrient-dense foods during the peri-transplant period. <u>There are no dietary restrictions imposed by the study protocol.</u>

A similar approach has been trialed successfully in myeloma patients <u>outside</u> the transplant setting. Because of the GI toxicities commonly experienced by patients who receive high-dose melphalan conditioning, we are conducting a feasibility pilot in this specialized population. We will explore how much of the delivered meals patients are able to eat, test our data collection mechanisms that would be used in a future randomized clinical trial (including food records, to be analysed collaboratively with the Fred Hutch Nutrition Assessment Shared Resource; NASR), and assess the magnitude of changes in gut microbiota and metabolome related to our trial diet compared with historical control patients.

A schedule of study activities is outlined in Appendix 1. Patients enrolled in the study will be seen by a clinical registered dietitian per standard practice via inpatient or outpatient policies which tailor the frequency of visits to acuity. Patients will also be followed by a research study registered dietitian weekly. Patients will be monitored clinically and by laboratories according to standard algorithms for approximately 1 month after HCT.

In addition to the 3 meals per day and snacks, delivered by Thistle from days -7 to +28, we will offer patients a list of additional plant-based supplement options in the form of a written guide. Logistics for the food deliveries are as follows:

- Deliveries will be made twice per week to the patients' home residence (whether that is their usual home or temporary housing during the peri-transplant period; *e.g.* SLU House).
- Meals are packaged in a cooler bag with ice packs to ensure temperature control during the transportation period.
- There is minimal meal preparation required: transfer to a microwaveable container followed by 2-3 minutes of heating is sufficient for most meals, or heating for similar duration in a skillet.
- If admitted to UW during the trial period, we will encourage caregivers to bring the delivered meals to the hospital, where they can be stored for short periods of time as any other meal brought from home would be.
- There is a 'default' weekly menu, but patients can access the online ordering system and make adjustments to the next week's deliveries before a defined cut-off (typically Thursday evening, when deliveries are made on Sunday and Wednesday evenings).
- Delivered meals will meet the Fred Hutchinson Cancer Center guidelines for food safety in immunosuppressed patients.

Thistle are a commercial company, providing home-delivered meals, operating throughout the US (a critical component for this pilot study as we move toward a randomized multicenter trial in the future). They offer a predominantly plant-based menu (in their commercially available meals, there is the option to add meats to some meals. Lunch and dinner meals contain an average of 450-650 calories and greater than 20g of plant protein from legumes, nuts & seeds, tempeh, tofu, and whole grains.

<u>Patients are not restricted or prevented from adding additional foods (within our standard food safety guidelines) by participating in the trial, nor are they prevented or restricted from standard nutritional</u>

Fred Hutch IRB
Approved
8/7/2024

support that may be offered during the course of transplant (e.g. tube feeding, parenteral nutrition in the case of severe gut toxicities).

Because the study intervention is the provision of food, we will not collect adverse events and severe adverse events unless they are possibly, probably, or definitely related to the ingestion of the plant-based whole foods or one of the data collection procedures. Medical complications that are attributed to the transplant procedure or myeloma will not be collected.

#### 5.1 Assessments

**Basic patient data:** Including demographics, prior treatments, co-morbidities to calculate HCT-CI will be collected by reviewing the medical record.

**Dietary assessments:** In collaboration with NASR, we will collect a 24-hr recall at 3 time points per patient (baseline, during neutropenic nadir, and at study end). We will encourage use of the multiday food record to record all meals eaten throughout, and completeness of the record will be assessed as an aspect of feasibility.

**Orientation to Thistle platform and Weekly Visit with Research Dietician:** The initial orientation will be in the form of a meeting with the study dietitian and will be accompanied by written/visual materials for how to access the Thistle platform. This can be done via Smartphone or computer. For patients without access to either, we will facilitate access during clinic follow up and also record this lack of access as a learning point throughout the pilot study. The weekly visits will occur between Day -7 to Day+28 period.
**Routine lab results and body weight measurements**: A Nutrition Focused Physical Exam will be performed on arrival by the clinical registered dietitian as per standard of care and again at the d28+/-7 day time point.

**Medical complications:** The medical record review will collect data related to oral intake, nutritional goals (from the clinical and research dietitians notes), diarrhea, and other gastrointestinal symptoms (from the medical notes). Reasons and dates for starting, stopping, and changing nutritional approaches will be documented (e.g., cessation of oral diet and introduction of nutritional support like tube feeds or TPN).

**Transplant outcomes:** We will assess admission to hospital from outpatient setting, infections, use of antibiotics, blood count recovery, organ function and biochemistry, cancer relapse, and death.

**Fecal microbiome:** Stool samples for microbiome assessment will be collected once prior to diet initiation and weekly throughout the study period. Stool samples will be processed by the SPL (aliquoted into at least 6 aliquots per sample). Sequencing will be performed using the external company Zymo Genomics. Metabolomics will be performed with the Northwest Metabolomics Center.

Fred Hutch IRB
Approved
8/7/2024

**Blood collection for metabolite and cell analysis:** 30 mL of blood will be collected at scheduled clinical blood draws once pre-transplant, and at day 28 (within +/-7 days). We will use these samples to profile the baseline and recovering immune system after transplantation using flow cytometry.

**Quality of life surveys:** Before diet initiation and an d28 +/- 7 days, patients will complete the EORTC QLQ-MY-20 and the QLQ-C30

**Initial patient survey:** At one of the initial visits, patients will be presented with a short initial survey to document GI symptoms and previous experience with a plant-based diet.

**Exit survey:** On d28 +/- 7 days, patients will be presented with a short 'exit survey' to formally document their opinion of the plant-based whole foods approach and gather qualitative data regarding intestinal symptoms.

#### 5.2 Study Endpoints

##### 5.2.1 Primary Endpoints

Feasibility and tolerability

- The proportion of approached patients who enroll
- Percentage of stool samples, blood samples and surveys collected
- Tolerability of the advised diet, based on qualitative end-study surveys and proportions of the delivered meals consumed.

##### 5.2.2 Secondary and Exploratory Endpoints

- Disease response at 28 days (IMWG disease-response criteria);[26]
- Microbiome diversity and domination with pathobionts (e.g. *Enterococcus*), the relationship of microbial diversity with food diversity as measured on the 3 x 24-hour recall assessments.
- Preservation of short-chain fatty acid-producing bacteria;
- Concentration of short-chain fatty acids in stool
- Immunophenotype d+ 28 (flow cytometry on blood samples);
- Quality of life outcomes (EORTC QLQ-MY-20, specifically tailored to people living with MM with the standard QLQ-C30).[27]
- We will trial indices for describing food intake e.g. the Healthy Eating Index, which can be derived from the 24-hr recall.[6]

### 6 BIOSTATISTICAL ANALYSIS

The analysis of the study will mainly be descriptive. The primary objectives will be analyzed as follows.

- The proportion of eligible patients who consent to enroll in the study will be computed based on the number approached and estimated with 95% confidence intervals (CI). Reasons for non-participation will be summarized.

Fred Hutch IRB
Approved
8/7/2024

- Feasibility of endpoint data collection will be measured by the percentage of stool samples, blood samples and patient surveys collected.
- Patient opinion/acceptability of the intervention will be gauged based on the end-of-study survey.

Greater than 50% for the feasibility endpoints will be considered successful.

Secondary endpoints including biologic measures of nutritional status, fecal microbiome and immune parameters will be assessed. Microbiome data will be used to provide preliminary data needed to design larger randomized controlled trials examining the impact of the intervention on the gut microbiome and metabolome.

Patient surveys will be scored according to the recommendations of the developers and reported descriptively.

### 7 RISKS AND DISCOMFORTS

Participation in this study involves receiving plant-based whole foods meals and <u>ideally, also consuming them</u> during the peri-transplantation period. Participants will be clearly told that there are no restrictions on other dietary intake as long as they follow our standard food safety guidelines.

The commercial company “Thistle” delivering the meals will have access to the study participants name, email address, physical delivery address, and contact phone number, but no other patient details will be shared with the company. This information will be given to Thistle when the deliveries are set up in their online system, with the patients specific informed consent.

The investigator must assure that participants’ anonymity will be strictly maintained and that their identities are protected from unauthorized parties. All research data will be kept in a locked file by the investigators. Study questionnaires and answers are confidential and will not be included in a patient’s medical record. Data from the questionnaires will be stored in a password-protected database within the institutional firewall.

**Known Potential Risks:**

<u>Risks of Stool Collection</u>
Stool collection can be embarrassing or messy for subjects.

<u>Risks of Blood Draw</u>
Blood samples will be collected by trained medical staff, and antiseptic procedures will be used for all blood draws. Most subjects will have blood drawn from their central venous catheter. Subjects having blood drawn from a vein may experience pain, lightheadedness, syncope, or bruising and/or swelling around the draw site. There is a possible risk of infection with blood draws from either the central venous catheter or from a vein.

<u>Risks of Patient Surveys</u>

The patient surveys are not designed to collect sensitive information, but subjects may be uncomfortable reporting gastrointestinal symptoms and dietary intake.

Unknown Risks
There may be unknown risks associated with the interventions for this study.

### 8 POTENTIAL BENEFITS

While we do not know if participants will benefit from participation in the study, it is possible that patients who are receiving prepared meals delivered to them directly will enjoy an improved nutritional status throughout the peri-transplant period. This may be linked with changes that are beneficial during transplant such as improved gut barrier function and reduced symptoms like diarrhea. We hope to use information from this study to design a larger study to evaluate the effects of a plant-based whole foods diet versus standard of care.

### 9 MONITORING AND QUALITY ASSURANCE

All dietitians have received specialty training and have the appropriate certifications and licenses. The study dietician will oversee the rest of the team. A list of suggested oral nutrition supplements will be provided.

All data collected will be kept in locked file cabinets and/or entered into a password-protected REDCap database using subject ID. The file linking subject ID to patient identity will be kept separately.

#### 9.1 Adverse Event Grading

All grade 3 or higher AEs according to the NCI Common Terminology Criteria for Adverse Events (CTCAE) Version 5.0 attributed to the meal delivery, consumption of a predominantly plant-based diet or collection of endpoint data that require intervention will be recorded. All serious, related, and unanticipated adverse events will be reported to the IRB in accordance with its policies, and at least annually as required by the IRB. Other adverse events and severe adverse events that are not attributable to study participation will not be recorded or reported.

Abnormal laboratory values for laboratory parameters specified in the study should not be recorded as an adverse event unless an intervention is required (repeat testing to confirm the abnormality is not considered intervention), the laboratory abnormality results in a serious adverse event or the adverse event results in study termination or interruption/discontinuation of study treatment.

Medical conditions present at screening are not adverse events and will not be recorded. Medical conditions present at baseline that worsen in intensity or frequency during the study period will be reported and recorded as adverse events if they are related to study participation and require intervention.

#### 9.2 Serious Adverse Event

An adverse event should be classified as an SAE if it meets one of the following criteria:

| | |
|---|---|
| Fatal | Adverse event results in death. |

Fred Hutch IRB
Approved
8/7/2024

| | |
|---|---|
| Life threatening: | The adverse events placed the subject at immediate risk of death. This classification did not apply to an adverse event that hypothetically might cause death if it were more severe. |
| Hospitalization: | It required or prolonged inpatient hospitalization. Hospitalizations for elective medical or surgical procedures or treatments planned before enrollment in the treatment plan or routine check-ups are not SAEs by this criterion. Admission to a palliative unit or hospice care facility is not considered to be a hospitalization. |
| Disabling/incapacitating | Resulted in a substantial and permanent disruption of the subject's ability to carry out normal life functions. |
| Congenital anomaly or birth defect: | An adverse outcome in a child or fetus of a subject exposed to the molecule or treatment plan regimen before conception or during pregnancy. |
| Medically significant: | The adverse event did not meet any of the above criteria, but could have jeopardized the subject and might have required medical or surgical intervention to prevent one of the outcomes listed above. |

#### 9.3 Monitoring and Recording Adverse Events

All reportable grade 3 or higher AEs will be assessed by the investigator or qualified designee and recorded in the CRFs. The investigator should attempt to establish a diagnosis of the event on the basis of signs, symptoms and/or other clinical information. In such cases, the diagnosis should be documented as the adverse event and/or serious adverse event and not described as the individual signs or symptoms. The following information should be recorded:

- Description of the adverse event using concise medical terminology
- Description as to whether or not the adverse event is serious, noting all criteria that apply
- The start date (date of adverse event onset)
- The stop date (date of adverse event resolution)
- The severity (grade) of the adverse event
- A description of the potential relatedness of the adverse event to study drug, a study procedure, or other causality
- The action taken due to the adverse event
- The outcome of the adverse event

#### 9.4 Grading Adverse Event Severity

The NCI Common Terminology Criteria for Adverse Events (CTCAE) Version 5.0 will be used for grading.

Clinical significance of AEs will be determined in the context of HCT and will take into account the expectedness of events.

Fred Hutch IRB
Approved
8/7/2024

#### 9.5 Attribution of an Adverse Event

Only grade 3 or higher AEs attributed to meal delivery, consumption of delivered meals or collection of endpoint data will be recorded. An AE is considered related if it is assessed as definitely, probably, or possibly related and unrelated if it is assessed as unlikely related or unrelated as defined below:

- Definite: The event follows a reasonable temporal sequence from exposure to the investigational agent, has been previously described in association with the investigational agent, and cannot reasonably be attributed to other factors such as the subject’s clinical state, other therapeutic interventions, or concomitant medications; AND the event disappears or improves with withdrawal of the investigational agent and/or re-appears on re-exposure (e.g., in the event of an infusion reaction).
- Probable: The event follows a reasonable temporal sequence from exposure to the investigational agent and has been previously described in association with the investigational agent OR cannot reasonably be attributed to other factors such as the subject’s clinical state, other therapeutic interventions, or concomitant medications.
- Possible: The event follows a reasonable temporal sequence from exposure to the investigational agent, but could be attributable to other factors such as the subject’s clinical state, other therapeutic interventions or concomitant medications.
- Unlikely: Toxicity is doubtfully related to the investigational agent(s). The event may be attributable to other factors such as the subject’s clinical state, other therapeutic interventions, or concomitant medications.
- Unrelated: The event is clearly related to other factors such as the subject’s clinical state, other therapeutic interventions, or concomitant medications.

#### 9.6 Adverse Event Recording Period

AEs with an onset date prior to the start of enrollment will not be recorded, except in the case of clinically significant worsening of the AE during the specified AE monitoring time frame.

#### 9.7 Reporting Protocol Deviations

A protocol deviation is any change, divergence, or departure from the study design or procedures defined in the protocol. Important protocol deviations are a subset of protocol deviations that may significantly impact the accuracy, and/or reliability of the study data or that may significantly affect a participant's rights, safety, or well-being. The principal investigator and personnel are responsible for identifying and reporting important deviations. Once important protocol deviations are identified, corrective actions are to be developed and implemented promptly. Protocol deviations must be sent to the IRB per their guidelines.

Fred Hutch IRB
Approved
8/7/2024

### 10 APPENDICES

I. **Schedule of Study Activities**
II. **Patient Initial survey**
III. **End of Study survey**
IV. **Multi-day food record sample document**
V. **QOL surveys**

Fred Hutch IRB
Approved
8/7/2024

#### APPENDIX 1: Schedule of Study Activities

| **Study Activity Timing** | **Enrollment** | **Baseline visit (may be concurrent with enrollment)[1]** | **Neutropenic nadir (d10+/-5)** | **d-7 to d+28[2]** | **Day 28 +/- 7 d** | **End of Study[3]** |
|---|---|---|---|---|---|---|
| Consent | X | | | | | |
| Orientation to the Thistle platform and ordering[4] | | X | | | | |
| Meals delivered | | | | 2 times a week | | |
| Visit with the study dietitian | | | | Weekly | | |
| Assessment of Outcomes[5] |  | ----------------------------------------- Continuous --------------------------------------- |  |  |  |  |
| ***Surveys*** | | | | | | |
| Formal 24-hr food recall | | X | X | | | X |
| Patient Initial survey [6] | | X | | | | |
| Multi-day food record | | X | | 3 times a week | | |
| QOL survey[7] | | X | | | | X |
| End of Study Survey | | | | | | X |
| ***Sample Collection*** | | | | | | |
| Stool Collection | X | | | Weekly | X | |
| Blood Collection | X | | | | X | |

[1] The baseline activities need to be completed prior to the first food delivery, but otherwise can happen any time post-enrolment.

[2] The meal delivery schedule is Sunday PM and Wednesday PM. There will be some variation to the timing to accommodate these delivery days. All assessments will have a window of +/- 3 days except Day +28 visit that will have a +/- 7 day window.

[3] Defined as discharge from the auto-transplant service, typically 1 month following autologous stem cell transplantation. Otherwise, will be marked as the completion of meal deliveries (~4 weeks post transplantation). Study participation may end before that time for other reasons, such as study withdrawal by either the participant, their provider, or the study's principal investigator.

[4] This will be in the form of a meeting with the study dietitian, and will be accompanied by written/visual materials for how to access the Thistle platform. This can be done via Smartphone or computer. For patients without access to either, we will facilitate access during clinic follow up and also record this lack of access as a learning point throughout the pilot study.

[5] Clinical monitoring via review of the electronic medical record will be continuous starting at time of consent and may continue indefinitely as needed for collection of health outcomes.

[6] See Appendix 2.

[7] See Appendix 5.

Fred Hutch IRB
Approved
8/7/2024

#### APPENDIX II: Patient Initial Survey

Please circle your responses to the following questions about yourself, your experiences with and attitudes towards nutrition, and recent gastrointestinal symptoms:

1. Have you ever received supplemental nutrition? (Circle all that apply)

   Yes, enterally (via a feeding tube) ..................................1
   Yes, parenterally (via a central line*) ..............................2
   Yes, via oral supplements (Ensure, Boost, etc.)...............3
   No ..........................................................................4
   *a central line is a Hickman, Groshong, or PICC line

2. Which statement best matches your feelings about the importance of nutrition during transplantation?

   Extremely important .....................................................1
   Important .....................................................................2
   Somewhat important ....................................................3
   Unimportant..................................................................4
   Do not know .................................................................5
   Other (please specify): ____________________________6

3. Have you ever eaten a plant-based diet in the past, or currently?

   No ☐
   Yes ☐

   If Yes:
   Current ☐
   Past ☐

4. In the past 7 days, how often did you have nausea – that is, a feeling like you could vomit?

   Never ........................................................................1
   Rarely.........................................................................2

Fred Hutch IRB
Approved
8/7/2024

Sometimes......................................................................3
Often ..............................................................................4
Always ............................................................................5

5. In the past 7 days, how often did you have a poor appetite?

Never ..............................................................................1
Rarely..............................................................................2
Sometimes......................................................................3
Often ..............................................................................4
Always ............................................................................5

6. In the past 7 days, how often did you throw up or vomit?

Never ..............................................................................1
One day ..........................................................................2
2-6 days ..........................................................................3
Once a day......................................................................4
More than once a day ....................................................5

7. In the past 7 days, how many days did you have loose or watery stools?

No days...........................................................................1
1 day ...............................................................................2
2 days .............................................................................3
3-5 days ..........................................................................4
6-7 days ..........................................................................5

8. In the past 7 days, how much did having loose or watery stools bother you?

Not at all .........................................................................1
A little bit........................................................................2
Somewhat ......................................................................3
Quite a bit.......................................................................4
Very much ......................................................................5

9. In the past 7 days, how often did you have belly pain?

Never ..............................................................................1
One day ..........................................................................2
2-6 days ..........................................................................3

Fred Hutch IRB
Approved
8/7/2024

Once a day............................................................................4
More than once a day ......................................................5

10. In the past 7 days, how much did your belly pain bother you?

Not at all ...............................................................................1
A little bit ..............................................................................2
Somewhat .............................................................................3
Quite a bit ..............................................................................4
Very much .............................................................................5

Fred Hutch IRB
Approved
8/7/2024

#### APPENDIX III: Patient End of Study Survey

1. Were you able to order the Thistle meals using the app or computer without assistance from the study team?

   Yes ☐

   No ☐

   If no: Please describe the help you needed, and how we can do this better in future.

2. Were there any problems with the delivery process?

   No ☐

   Yes ☐

   If yes: Please describe the issues so we can fix this in future.

3. Did you get admitted to hospital during the study period?

   No ☐

   Yes ☐

   If yes: were you able to continue eating the Thistle meals during the admission?

   Yes ☐

   No ☐

If no: please describe the barriers to continuing to eat the meals (e.g. GI symptoms like nausea, unable to get them in hospital, lack of support to heat the up etc).

Fred Hutch IRB
Approved
8/7/2024

4. In the past 7 days, how often did you have nausea – that is, a feeling like you could vomit?

Never ..................................................................1
Rarely..................................................................2
Sometimes..........................................................3
Often ..................................................................4
Always ................................................................5

5. In the past 7 days, how often did you have a poor appetite?

Never ..................................................................1
Rarely..................................................................2
Sometimes..........................................................3
Often ..................................................................4
Always ................................................................5

6. In the past 7 days, how often did you throw up or vomit?

Never ..................................................................1
One day ..............................................................2
2-6 days ..............................................................3
Once a day..........................................................4
More than once a day .......................................5

7. In the past 7 days, how many days did you have loose or watery stools?

No days...............................................................1
1 day ...................................................................2
2 days .................................................................3
3-5 days ..............................................................4
6-7 days ..............................................................5

8. In the past 7 days, how much did having loose or watery stools bother you?

Not at all.............................................................1
A little bit...........................................................2
Somewhat ...........................................................3
Quite a bit...........................................................4
Very much ..........................................................5

Fred Hutch IRB
Approved
8/7/2024

9. In the past 7 days, how often did you have belly pain?

Never ..................................................................1
One day ..............................................................2
2-6 days ..............................................................3
Once a day ..........................................................4
More than once a day .........................................5

10. In the past 7 days, how much did your belly pain bother you?

Not at all .............................................................1
A little bit ............................................................2
Somewhat ...........................................................3
Quite a bit ............................................................4
Very much ...........................................................5

Fred Hutch IRB
Approved
8/7/2024

#### APPENDIX IV: Multi-Day Food Record Sample

**Thistle Meal Study Daily Checklist**

| DAY ______________ DATE____________________ | | | | |
|---|---|---|---|---|
| | | **Did you eat it all?** | | |
| | **Thistle Meal** [write down the full name of the Thistle food/meal eaten] | **Yes (√)** | **No (estimate % eaten)** | **Notes** |
| *Breakfast* | | ☐ | | |
| *Lunch* | | ☐ | | |
| *Dinner* | | ☐ | | |
| *Snack* | | ☐ | | |

Did you eat or drink anything (including coffee/tea) that was not provided? ☐ Yes ☐ No

If yes, please record any non-study foods, condiments and/or beverages you had on this day.

| Meal (B'fast, Lunch, Dinner, Snack) | Food/condiment/beverage | Brand/ Restaurant name | Preparation | Amount |
|---|---|---|---|---|

Fred Hutch IRB
Approved
8/7/2024

#### APPENDIX V: EORTC Survey Samples

### EORTC QLQ-MY20

Patients sometimes report that they have the following symptoms or problems. Please indicate the extent to which you have experienced these symptoms or problems <u>during the past week</u>. Please answer by circling the number that best applies to you.

| **During the past week:** | | **Not at All** | **A Little** | **Quite a Bit** | **Very Much** |
|---|---|---|---|---|---|
| 31. | Have you had bone aches or pain? | 1 | 2 | 3 | 4 |
| 32. | Have you had pain in your back? | 1 | 2 | 3 | 4 |
| 33. | Have you had pain in your hip? | 1 | 2 | 3 | 4 |
| 34. | Have you had pain in your arm or shoulder? | 1 | 2 | 3 | 4 |
| 35. | Have you had pain in your chest? | 1 | 2 | 3 | 4 |
| 36. | If you had pain did it increase with activity? | 1 | 2 | 3 | 4 |
| 37. | Did you feel drowsy? | 1 | 2 | 3 | 4 |
| 38. | Did you feel thirsty? | 1 | 2 | 3 | 4 |
| 39. | Have you felt ill? | 1 | 2 | 3 | 4 |
| 40. | Have you had a dry mouth? | 1 | 2 | 3 | 4 |
| 41. | Have you lost any hair? | 1 | 2 | 3 | 4 |
| 42. | Answer this question only if you lost any hair: Were you upset by the loss of your hair? | 1 | 2 | 3 | 4 |
| 43. | Did you have tingling hands or feet? | 1 | 2 | 3 | 4 |
| 44. | Did you feel restless or agitated? | 1 | 2 | 3 | 4 |
| 45. | Have you had acid indigestion or heartburn? | 1 | 2 | 3 | 4 |
| 46. | Have you had burning or sore eyes? | 1 | 2 | 3 | 4 |

Fred Hutch IRB
Approved
8/7/2024

| **During the past week:** | **Not at All** | **A Little** | **Quite a Bit** | **Very Much** |
|---|---|---|---|---|
| 47. Have you felt physically less attractive as a result of your disease or treatment? | 1 | 2 | 3 | 4 |
| 48. Have you been thinking about your illness? | 1 | 2 | 3 | 4 |
| 49. Have you been worried about dying? | 1 | 2 | 3 | 4 |
| 50. Have you worried about your health in the future? | 1 | 2 | 3 | 4 |

#### EORTC QLQ-30 (Version 3)

We are interested in some things about you and your health. Please answer all of the questions yourself by circling the number that best applies to you. There are no "right" or "wrong" answers. The information that you provide will remain strictly confidential.

Please fill in your initials: ☐☐☐☐
Your birthdate (Day, Month, Year): ☐☐☐☐☐☐☐☐
Today's date (Day, Month, Year): 31 ☐☐☐☐☐☐☐☐

| | **Not at All** | **A Little** | **Quite a Bit** | **Very Much** |
|---|---|---|---|---|
| 1. Do you have any trouble doing strenuous activities, like carrying a heavy shopping bag or a suitcase? | 1 | 2 | 3 | 4 |
| 2. Do you have any trouble taking a <u>long</u> walk? | 1 | 2 | 3 | 4 |
| 3. Do you have any trouble taking a <u>short</u> walk outside of the house? | 1 | 2 | 3 | 4 |
| 4. Do you need to stay in bed or a chair during the day? | 1 | 2 | 3 | 4 |
| 5. Do you need help with eating, dressing, washing yourself or using the toilet? | 1 | 2 | 3 | 4 |

| **During the past week:** | **Not at All** | **A Little** | **Quite a Bit** | **Very Much** |
|---|---|---|---|---|
| 6. Were you limited in doing either your work or other daily activities? | 1 | 2 | 3 | 4 |

Fred Hutch IRB
Approved
8/7/2024

| | | | | |
|---|---|---|---|---|
| 7. Were you limited in pursuing your hobbies or other leisure time activities? | 1 | 2 | 3 | 4 |
| 8. Were you short of breath? | 1 | 2 | 3 | 4 |
| 9. Have you had pain? | 1 | 2 | 3 | 4 |
| 10. Did you need to rest? | 1 | 2 | 3 | 4 |
| 11. Have you had trouble sleeping? | 1 | 2 | 3 | 4 |
| 12. Have you felt weak? | 1 | 2 | 3 | 4 |
| 13. Have you lacked appetite? | 1 | 2 | 3 | 4 |
| 14. Have you felt nauseated? | 1 | 2 | 3 | 4 |
| 15. Have you vomited? | 1 | 2 | 3 | 4 |
| 16. Have you been constipated? | 1 | 2 | 3 | 4 |

| **During the past week:** | **Not at All** | **A Little** | **Quite a Bit** | **Very Much** |
|---|---|---|---|---|
| 17. Have you had diarrhea? | 1 | 2 | 3 | 4 |
| 18. Were you tired? | 1 | 2 | 3 | 4 |
| 19. Did pain interfere with your daily activities? | 1 | 2 | 3 | 4 |
| 20. Have you had difficulty in concentrating on things, like reading a newspaper or watching television? | 1 | 2 | 3 | 4 |
| 21. Did you feel tense? | 1 | 2 | 3 | 4 |
| 22. Did you worry? | 1 | 2 | 3 | 4 |
| 23. Did you feel irritable? | 1 | 2 | 3 | 4 |
| 24. Did you feel depressed? | 1 | 2 | 3 | 4 |
| 25. Have you had difficulty remembering things? | 1 | 2 | 3 | 4 |
| 26. Has your physical condition or medical treatment interfered with your family life? | 1 | 2 | 3 | 4 |

| | | | | |
|---|---|---|---|---|
| 27. Has your physical condition or medical treatment interfered with your <u>social</u> activities? | 1 | 2 | 3 | 4 |
| 28. Has your physical condition or medical treatment caused you financial difficulties? | 1 | 2 | 3 | 4 |

**For the following questions please circle the number between 1 and 7 that best applies to you**

29. How would you rate your overall <u>health</u> during the past week?

| 1 | 2 | 3 | 4 | 5 | 6 | 7 |
|---|---|---|---|---|---|---|
| Very poor | | | | | | Excellent |

30. How would you rate your overall <u>quality of life</u> during the past week?

| 1 | 2 | 3 | 4 | 5 | 6 | 7 |
|---|---|---|---|---|---|---|
| Very poor | | | | | | Excellent |
