## Supplementary methods for "Plant-based whole-food diets are feasible during autologous stem cell transplantation and are associated with dose-dependent microbiome modulation: Results from a pilot clinical trial"

*Shotgun metagenomic sequencing*

Shotgun Metagenomic Sequencing was performed by Zymo Research (Irvine, CA, USA) at a sequencing depth of 10 million paired-end 2x250 reads. In short, DNA extraction was performed on frozen fecal samples using a combination of ZymoBIOMICS-96 MagBead DNA Kit, ZymoBIOMICS DNA Miniprep Kit, and/or ZymoBIOMICS DNA Microprep Kit. According to the manufacturer’s instructions, library preps using the Illumina DNA Library Prep Kit (Illumina, San Diego, CA, USA) with dual-index 10 bp barcodes and Nextera adapters were pooled in equal abundance and quantified by qPCR and TapeStation (Agilent Technologies, Santa Clara, CA, USA). Pooled libraries were sequenced on the NovaSeq (Illumina, San Diego, CA, USA) against a ZymoBIOMICS Microbial Community Standard (Zymo Research) positive control and a blank prep negative control.

Low-quality fractions and adapters, host-derived reads, and low-diversity reads were removed with Trimmomatic-0.22,^1^ Kraken2,^2^ and, sdust (<https://github.com/lh2/sdust>), respectively. DIAMOND sequencing aligner^3^ assigned antimicrobial resistance and virulence gene factor identification by using NCBI repository-curated reference databases. Sourmash^4^ was used to profile microbial composition against a GTDB species representative database (RS207) and viral, protozoa, and fungi identification against GenBank databases (v.2022.03). These genomes were mapped and quantified based on counts to their corresponding reads via sourmash, using BWA-MEM. QIIME^5^ was used for alpha- and beta-diversity analysis and HUMAaN2^6^ was used for functional profiling.

*Metabolomics*

Fecal SCFA were quantified using LC-MS/MS as described.^7^ Briefly, dried fecal samples were extracted using methanol, then centrifuged, and the clear supernatants collected. For derivatization, 40 µL of each supernatant was mixed with 20 µL of 200 mM 3-nitrophenylhydrazine in 50% aqueous acetonitrile and 20 µL of 120 mM N-(3-dimethylaminopropyl)-N’-ethylcarbodiimide in 50% aqueous acetonitrile with 6% pyridine. The mixture was incubated at 40°C for 30 minutes. After reaction, the mixture was diluted to 2 mL with 10% aqueous acetonitrile.  A 500 µL aliquot was mixed with 500 µL of an isotope-labeled IS mixture, and a 20 µL aliquot was injected for LC-MS/MS on a Waters Acquity I-class UPLC coupled to a Waters Xevo TQS-micro MS. Chromatographic separations were performed on a Waters BEH C18 (2.1 x 50 mm, 1.7 µm) UPLC column. Fecal samples were dried using an Eppendorf Vacufuge vacuum concentrator. Fecal data were normalized using the mass of the dried sample, taken prior to extraction.

*Statistics*

To test for global differences in community structure, a permutational multivariate analysis of variance (PERMANOVA) was conducted using the *adonis/adonis2* functions in the *vegan* package. PERMANOVA models were fit to distance matrices (e.g., Bray–Curtis or Jaccard dissimilarity) and included covariates (time point and meal consumption status) as noted in the figure legends, with significance assessed using permutation-based P values (typically 999 or more permutations). Model terms were interpreted as the proportion of variance in the distance matrix explained by each factor (R²), recognizing that PERMANOVA is sensitive to differences in both centroids and multivariate dispersion.

Group differences (time or consumption status) were evaluated using unpaired nonparametric Wilcoxon tests. Where paired samples were available, within-subject changes over time were assessed using paired testing.

EORTC QLQ-MY20 and QLQ-C30 outcomes were summarized using median and interquartile range, and analysis was performed in Microsoft Excel. All analyses shown in Figures 1-3 were performed in R/RStudio (version 2025.05.1+513), using base functions and the *vegan* package for community ecology analyses.

*Data sharing*

Shotgun metagenomic sequencing data and metabolomics are available at dbGaP (accession: *will be made available by the time of publication*) and the National Metabolomics Data Repository (NMDR; accession: *will be made available by the time of publication*).
